# Genetic Ancestry and Somatic Mutations in Lung Adenocarcinoma: Insights from Real-World Clinico-Genomic Data

**DOI:** 10.1101/2024.04.24.24306316

**Authors:** Brooke Rhead, Yannick Pouliot, Justin Guinney, Francisco M. De La Vega

## Abstract

**Background:** Lung cancer presents a significant global health challenge, with disparities in incidence and outcomes across races and ethnicities. These disparities underscore the need to explore the molecular landscapes of lung cancer in relation to ancestry. Here, we leverage data from a real-world clinico-genomic database to discover associations between molecular profiles and genetic ancestry or race/ethnicity categories.

**Methods:** We utilized data from a cohort of 13,196 primarily late-stage non-small cell lung adenocarcinoma (LUAD) patients, sequenced with the Tempus xT NGS 648-gene panel, of which normal tissue was also sequenced for 6,520 cases. Genetic ancestry proportions were estimated using ancestry informative markers. Race and ethnicity categories were imputed using an ancestry-backed method, resulting in the assignment of 568 Hispanic/Latino, 892 non-Hispanic (NH) Asian, 1,581 NH Black, and 10,063 NH White individuals. Multiple imputation addressed missing data on smoking status. Logistic regression models assessed associations between ancestry proportions and somatic variants in 23 LUAD-related genes, adjusting for a false discovery rate of 5%. Analyzed mutations included copy number alterations, gene fusions, protein-altering SNVs and indels, and actionable or predicted driver mutations.

**Results:** Our analysis confirmed previously reported associations, such as a positive correlation between East Asian (EAS) ancestry and *EGFR* (OR per doubling ancestry=1.1) and a negative correlation with *KRAS* driver mutations (OR=0.96), while European ancestry exhibited the opposite relationship (OR=0.93 and 1.08, correspondingly; all p<0.0001). We also verified a positive association with *EGFR* driver mutations (OR=2) and a negative one with *KRAS* (OR=0.46; p<0.001) among Hispanic/Latino patients and American Indigenous (AMR) genetic ancestry (OR=1.03 and 0.97, correspondingly; p<0.05). Novel associations were identified between African (AFR) and South Asian (SAS) ancestries and LUAD genes. Some associations are explained by differences in smoking status (e.g., *ATM* and *ALK* fusions), while others persist even after adjusting for smoking (e.g., *EGFR*, *KRAS*, and *CDKN2A* copy-number alterations). Notably, we identified a positive association between EAS ancestry and the imputed NH Asian category with driver mutations in *CTNNB1* (OR=1.05 and 2.2, respectively; p<0.01), independent of smoking. These mutations are rare in NH White patients (2.4%) but are more prevalent in never-smoker NH Asian patients with predominant EAS ancestry (8.5%).

**Conclusion:** This study underscores the value of clinico-genomic databases in revealing associations between LUAD mutational profiles and genetic ancestry, shedding light on lung cancer disparities. Identification of a previously unappreciated association between EAS with *CTNNB1*, a potential biomarker for spindle assembly checkpoint kinase (*TTK*) inhibitors effectiveness and prognosis in LUAD, emphasizes the value of studying diverse populations in cancer research, paving the way for more equitable lung cancer treatments.

## Introduction

Lung cancer is a multifaceted disease, shaped by a mix of environmental exposures and genetic predispositions.^1^ While smoking is the foremost causal factor in lung cancer development, decreasing rates of smoking and associated mortality^2^ have highlighted the rising incidence of lung cancer in never-smokers.^3^ This shift accentuates the need to explore lung cancer’s contributing factors beyond smoking.

Non-small cell lung adenocarcinoma (LUAD), the most common lung cancer subtype,^4^ is distinguished by unique molecular and epidemiological characteristics. LUAD features diverse genetic alterations, such as mutations in *EGFR*, *KRAS*, and *ALK* genes,^4^ which have been reported to differ across race and ethnicity (R/E) groups.^5–10^ The incidence and mortality rates of LUAD also vary markedly among different R/E and regions.^11–13^

Disparities in lung cancer incidence and outcomes across R/E and gender suggests a complex interaction between genetic vulnerability and environmental factors. For instance, never-smoker women of East Asian descent have a higher prevalence of LUAD with *EGFR* mutations.^3^ Black men face disproportionately higher lung cancer rates and worse outcomes than their White counterparts, suggesting that socio-economic factors, healthcare access, and potential genetic differences contribute to lung cancer disparities.^9,12^ The distinct molecular profiles of lung cancer in Hispanic/Latino populations, such as the higher prevalence of *KRAS* mutations,^6^ further emphasize the importance of examining lung cancer’s molecular landscapes in the context of genetic ancestry and race/ethnicity to discover new insights for targeted treatments and personalized care.

Historically, the exploration of lung cancer’s molecular profiles in relation to genetic ancestry and R/E categories have been hampered by limited diversity in research cohorts, small sample sizes with minimal minority representation,^14^ and reliance on broad and overlapping US federal R/E categories.^15^ These categories, which group genetically and geographically diverse populations together (e.g. conflate East and South Asians in “Asian”), and obscure the potential range of genetic admixtures (e.g. in Blacks and Hispanic or Latinos), limit the understanding of lung cancer’s complexities.^16,17^

Real-world clinical genomics databases that aggregate de-identified data from patients undergoing clinical testing offer a valuable resource for overcoming these limitations.^18–20^ Such databases are growing rapidly, reflecting the increasing adoption of tumor profiling and liquid biopsies in treatment guidelines.^19^ Despite healthcare access disparities, the representation of minority groups in these databases has improved,^21,22^ providing a rich source of multimodal molecular data for investigating molecular associations with race, genetic ancestry, and other clinical factors.

However, challenges such as significant missingness in R/E data^21,23^ and gaps in clinical data, like smoking history,^24^ persist. To bridge these gaps, our study leverages the Tempus clinico-genomic database to explore the associations between LUAD molecular profiles and genetic ancestry or R/E categories, inferring continental-level genetic ancestry from molecular data.

Our approach has revealed both known and novel associations between somatic mutations and genetic ancestry and R/E groups. Notably, we uncovered population-based associations with biomarkers of drug effectiveness and prognosis in LUAD, demonstrating the importance of studying diverse populations to identify new therapeutic strategies and insights that may help address healthcare disparities.

## Methods

### Patient cohort

We obtained records for 13,196 cancer patients diagnosed with LUAD from the de-identified Tempus clinico-genomic database, which includes genomic and clinical data from cancer patients that underwent tumor profiling using the Tempus xT assay as part of their healthcare. Briefly, Tempus xT is a targeted, tumor-normal-matched DNA panel that detects single-nucleotide variants, insertions and/or deletions, and copy number variants in 648 genes, as well as chromosomal rearrangements in 22 genes with high sensitivity and specificity.^25,26^ Selection criteria included tumor profiling with the Tempus xT assay (v2-v4) from 2018 to 2022. For patients with multiple independent test results, we selected the results corresponding to the first collection date.

### Genetic ancestry estimation

We determined proportions of continental genetic ancestry employing a supervised variant of the ADMIXTURE algorithm for global genetic ancestry inference,^27^ following methodologies outlined in prior research.^22,28^ We estimated ancestry proportions across five major super-populations—Africa (AFR), American Indigenous (AMR), East Asia (EAS), Europe (EUR), and South Asia (SAS)—utilizing a custom set of 654 ancestry informative markers (AIMs) previously identified in the targeted sequencing regions of the Tempus xT assay.^22^ Reference allele frequencies for these AIMs were derived from the 1,000 Genomes Project,^29^ the Human Genome Diversity Project,^3033^ and the Simons Genome Diversity Project^31^ databases. Specifically for the AMR super-population, we omitted the admixed “AMR” group from the 1,000 Genomes Project, opting instead for allele frequencies of Native American individuals from the alternate sources to enhance accuracy in reflecting American Indigenous population similarities.

### Imputation of race and ethnicity

The categories of race and ethnicity in real-world data (RWD) follow the guidelines set by the US Office of Management and Budget.^15^ Yet, these classifications can complicate analyses due to the separate questions for race and ethnicity, leading to the practical approach of flattening these categories into distinct, non-overlapping categories used in this study:^32^ Hispanic or Latino, non-Hispanic (NH) Asian, NH Black, and NH White. We imputed these race and ethnicity categories based on genetic ancestry proportions, utilizing a boosted logistic regression machine learning algorithm as outlined in earlier research.^33^ Individuals without at least a 50% probability of belonging to one of the race/ethnicity groups were categorized as “No Call” (0.7%). Prior publications have validated the accuracy of this approach using data from the Tempus clinico-genomic database (correct rate of 96% and weighted error of 0.9%).^33^

### Statistical analyses

Somatic mutations in genes previously associated with LUAD^34–36, 37–39^ present on the Tempus xT gene panel with a mutation in at least one percent of patients in the selected cohort were tested for association with genetic ancestry proportions and race and ethnicity categories. Five mutation types were tested separately: protein-altering SNVs and indels, gene fusions, SCNAs, mutations with an OncoKB classification for any cancer type as therapeutic level one or two or resistance level one,^3740^ and LUAD driver mutations as predicted by the boostDM algorithm.^35^

A directed acyclic graph (DAG) helped us to visualize the confounders and causative factors affecting LUAD incidence, particularly the relationships among R/E or genetic ancestry, smoking, social determinants of health (SDOH) and other environmental exposures, genetic predispositions, and somatic mutations (Supplementary Figure 1). The DAG positions smoking as a key mediator in the pathway from race/ethnicity to somatic mutations, indicating that the relationship between genetic ancestry and mutation rates is modulated, in part, by smoking behaviors. Associations were determined using likelihood ratio tests (LRTs) comparing full and nested logistic regression models. Three analyses were performed for each gene and mutation type: (1) univariable analysis, with the full model consisting of an indicator for the presence of somatic mutations in the gene as the dependent variable and either genetic ancestry proportions or imputed race and ethnicity category as the independent variables, and an intercept-only nested model; (2) complete case analysis adjusted for smoking status, with the same models as in the univariable analysis but with smoking status included as an additional independent variable in both the full and nested models, with only patients with known smoking status included; and (3) multiple imputation analysis adjusted for smoking status, using the same models as complete case analysis but including all patients and utilizing multiple imputed values for smoking status and pooling results (see below). Compositional data analysis methods were used to enable inclusion of all five genetic ancestry proportions in the same models.

Specifically, genetic ancestry proportions were transformed into an isometric log ratio representation using the *pivotCoord* function in the *robCompositions* R package.^38^ Models including imputed race and ethnicity categories used NH White as the reference category. For each mutation type, LRT p-values were corrected for the number of genes tested using the Benjamini-Hochberg method to control the false discovery rate at 5%. For any association where the corrected LRT p-value was <0.05, all logistic regression p-values <0.05 for a specific genetic ancestry proportion or imputed race and ethnicity group from the full model were considered associated.

### Multiple imputation

Multivariate imputation by chained equations (MICE) was performed in R using the *mice* package.^39^ Variables included as predictors in imputation models were smoking status, gender, age quartile at collection of tumor specimen, U.S. census division of patients’ home address state derived from 3-digit Zip Code (Pacific, Mountain, West North Central, West South Central, East North Central, East South Central, New England, Middle Atlantic, or South Atlantic), genetic ancestry proportions transformed to isometric log ratio pivot coordinates, tumor grade, cancer stage, tumor mutational burden (TMB, in mutations/megabase), assay version, and indicators for the presence of mutations in the genes tested: (1) actionable mutations in *ATM, BRAF, CTNNB1, EGFR, KRAS, NF1, PIK3CA, RBM10, STK11,* and *TP53*; (2) *CDKN2A, EGFR, ERBB2, KRAS,* and *MET* SCNAs; and (3) gene fusions with *ALK* (Supplemental Figure 2). The following methods were used for variables with missingness: smoking status (46% missing), logistic regression; age quartile at collection of tumor specimen (19% missing), multinomial regression; U.S. census division (19% missing), random forests; tumor grade (64% missing), multinomial regression; cancer stage (32% missing), multinomial regression; TMB (0.05% missing), predictive mean matching. Ten datasets were imputed, each with 20 iterations of the MICE algorithm. Plots of the mean and standard deviation of each variable with missingness were examined to assess convergence of the MICE algorithm, and distributions of imputed values were compared to measured values to assess the quality of imputations. Likelihood ratio test statistics from the imputed datasets were combined using the *D3* function in the *mice* package,^41^ and logistic regression test statistics were combined using the *pool* function in the *mice* package using the “Rubin 1987” pooling rule.^40^ We observed that MICE iterations converge well in our data (Supplementary Figure 4), that the multiply imputed categorical variables align with observed data (Supplementary Figure 5), and that the multiply imputed smoking status is distributed similarly across R/E categories, albeit with a tendency toward over-imputation of current/former smoking status in Hispanic/Latino and NH Asian (Supplementary Figure 6).

## Results

### Patient characteristics

Our study examined a cohort of LUAD patients totaling 13,196 individuals. Patients were categorized by imputed R/E into Hispanic or Latino (N=568), NH Asian (N=891), NH Black (N=1,581), and NH White (N=10,063), with 93 patients not included in any of these categories (**Table 1**). While most patients have majority European genetic ancestry (EUR), there is also a substantial number of patients with African (AFR), American Indigenous (AMR) and East Asian (EAS) ancestries, while patients of South Asian ancestry (SAS) are less represented in our cohort (**Figure 1**). We also observed a conflation of EAS and SAS ancestries in the NH Asian category—a well-known challenge. A notable majority of patients (75%) were former or current smokers, especially within the NH Black (84%) and NH White (79%) groups. Ages at specimen collection and diagnosis were, on average, in the mid-to late-sixties across groups, with minor variations across R/E. Gender distribution maintained a balance across groups, albeit with a modest female majority overall in the cohort. In this study, we also analyzed a sub-cohort of patients that had paired tumor and normal (T/N) specimens sequenced (Supplementary Table 1).

**Figure 1.**
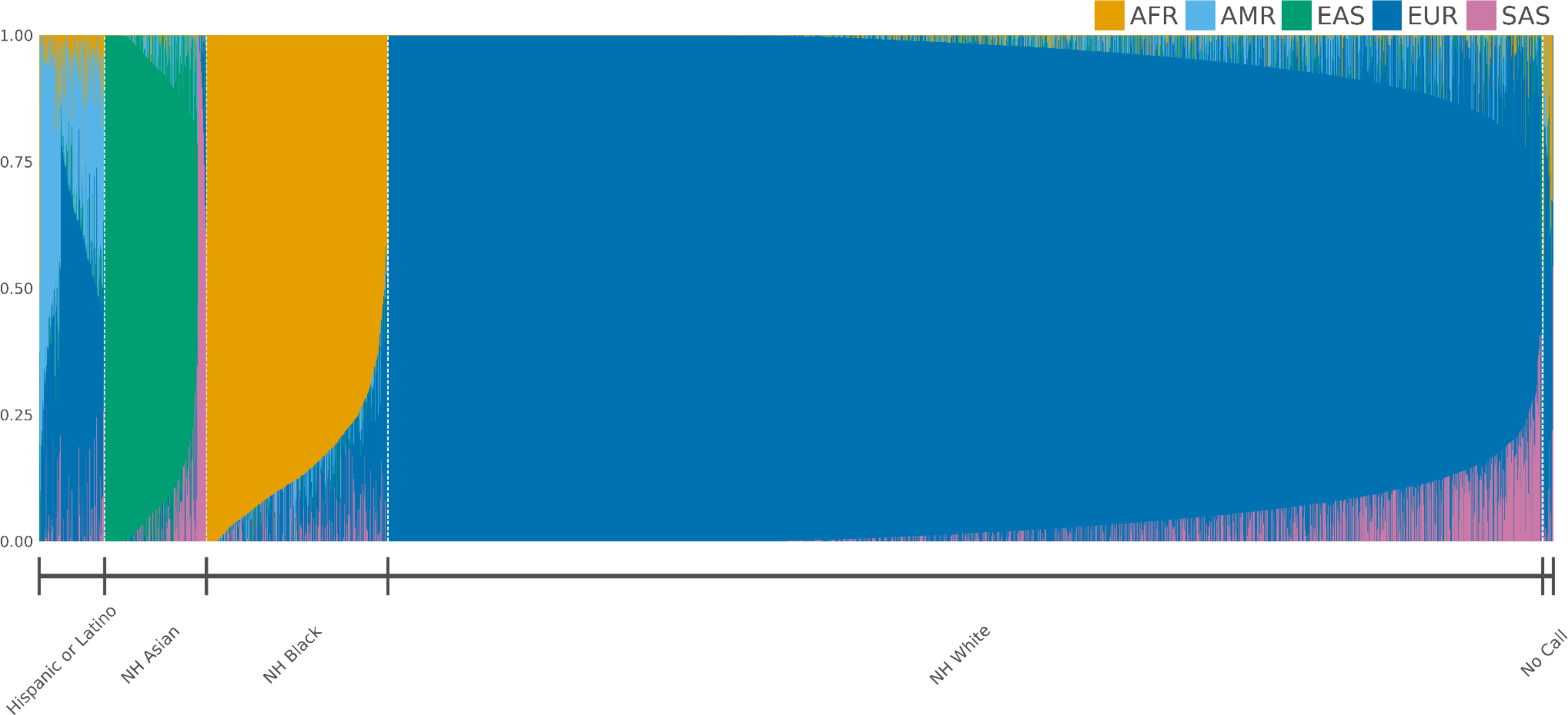
Ancestry proportions for patients in cohort by imputed race and ethnicity. Vertical bars represent the global continental ancestry admixture proportions for each patient, color-coded according to the scale provided in the legend. Patients are categorized by imputed race and ethnicity on the x-axis and are further sorted within each group by increasing values of their predominant continental ancestry.

**Table 1.**
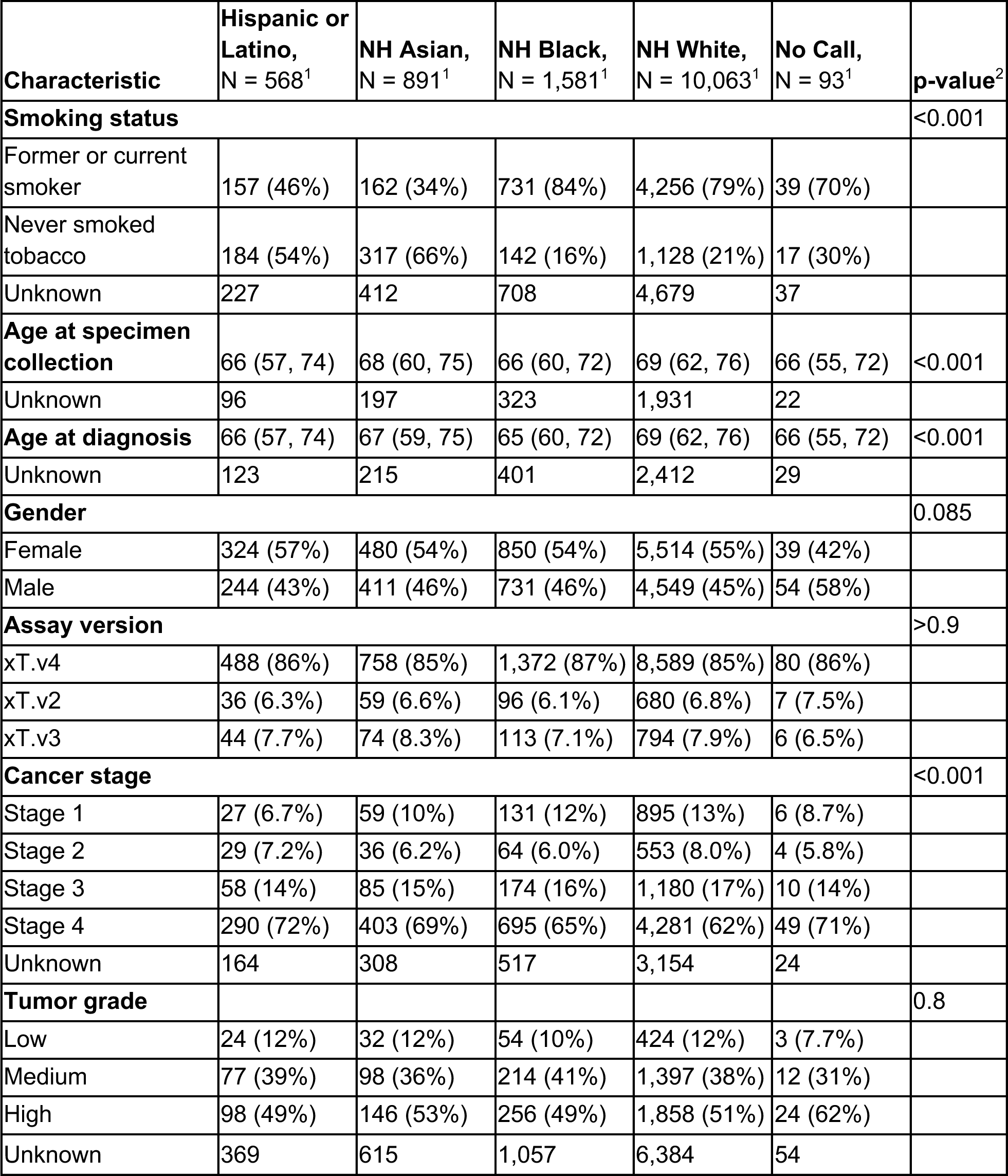
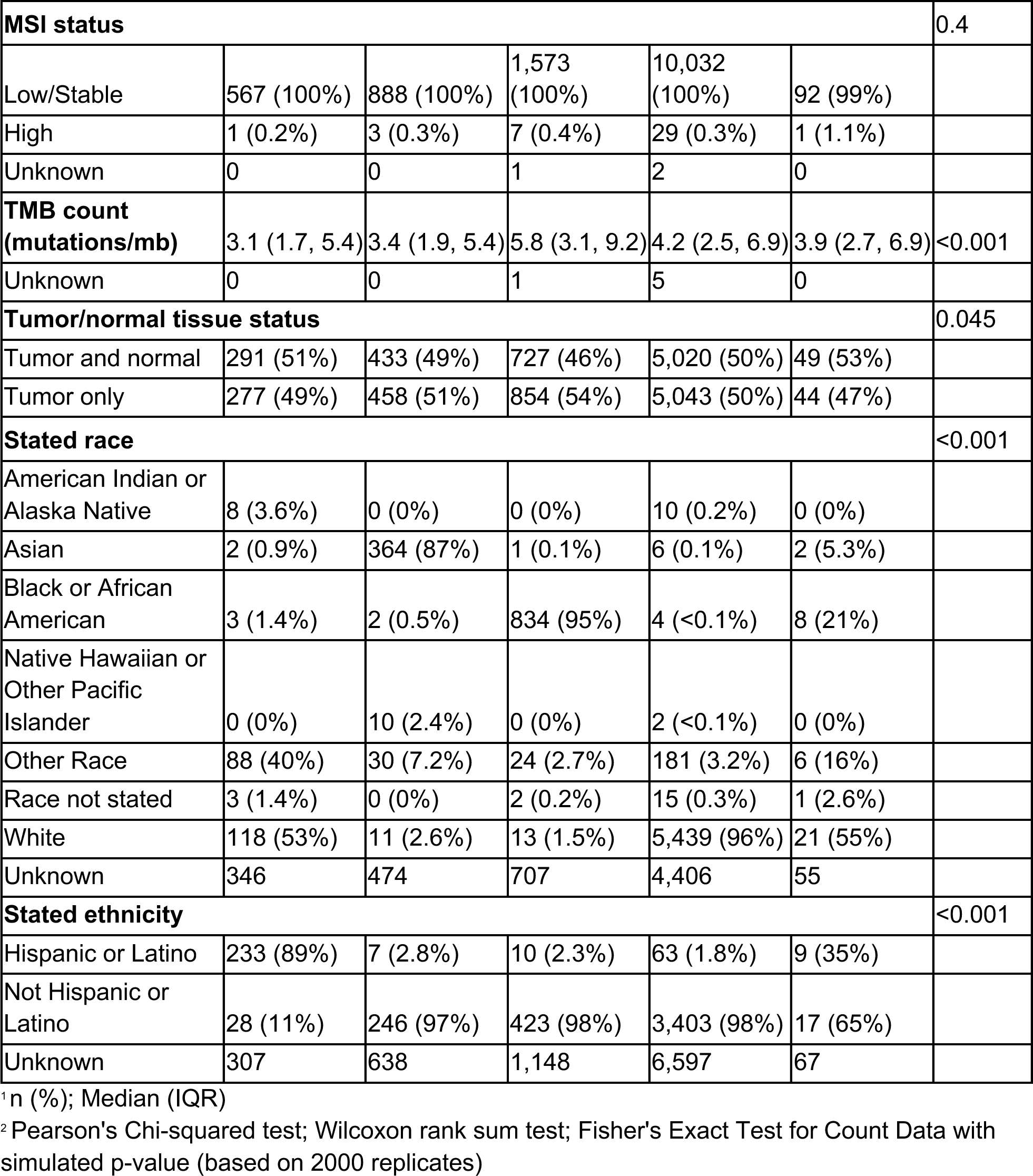
Cohort characteristics by imputed race and ethnicity category. Last column shows the results of statistical tests (as defined in footnote) for differences of the characteristics by imputed race and ethnicity.

Smoking status, age at specimen collection, and age at diagnosis all varied significantly across R/E categories (p<0.001), suggesting potential differences in disease exposures and disease course prior to sequencing (**Table 1, Supplementary Figure 1**). Of patients with known cancer stage, 63% had stage 4 cancer, consistent with expected utilization of tumor profiling in cancer care. This percentage was higher in the Hispanic or Latino (72%) and NH Asian (69%) categories (p<0.001), pointing to a higher prevalence of advanced disease at sequencing. TMB count also varied by R/E (p<0.005), with NH Black patients displaying a higher TMB count in the complete cohort (encompassing tumor only, TO, plus tumor-normal matched, T/N, sequencing modalities). However, TMB count in T/N cases, where misclassifications of germline variants as somatic is avoided, shows Hispanic/Latino and NH Asian exhibiting a lower burden (p<0.001; Supplementary Table 1).

Compared to never smokers, former or current smokers varied considerably by R/E, stated race, and stated ethnicity p<0.001), more male (48% vs. 32%), and had an earlier cancer stage, higher tumor grade, and higher TMB (average 5.0 vs. 2.3) (Supplementary Tables 2 and 3). There were no R/E differences in age at collection or diagnosis, assay version, microsatellite instability (MSI) status, or availability of normal tissue for sequencing. Compared to patients without available smoking status, patients whose smoking status was known varied by R/E (p=0.019) and stated race (p=0.01), were younger at time of specimen collection and at diagnosis (mean 67 vs. 71 in both cases; p<0.001), female (56% vs. 53% p<0.001)), more likely to be sequenced on an earlier xT assay version (82% xT.v4 vs. 90%; p<0.001)), had a slightly lower TMB count (4.2 on average vs. 4.6; p<0.001)), and were more likely to have normal tissue available for sequencing (54% vs. 44%). There was no difference in cancer stage, tumor grade, MSI status, or stated ethnicity.

We observed that smoking correlates with TMB, gender, and tumor grade (Supplementary Figure 2A) and mutations in genes such as *KRAS*, *EGFR*, and *ALK* fusions (Supplementary Figure 2B; Supplementary Figure 3A), as previously reported in the literature.^4^ Moreover, smoking status was associated with R/E (p<0.001; see **Table 1**) and affects frequency of somatic mutations in genes (Supplementary Figure 3B).

### Associations between genetic ancestry and somatic mutations

We tested for associations between continental genetic ancestry proportions and somatic alterations in genes with known oncogenic properties in LUAD (Supplementary Table 4, see Methods for selection criteria).

A univariable analysis revealed associations between genetic ancestry and somatic mutation patterns in several LUAD genes (**Table 2**). We confirmed a previously reported association between EAS with an increased frequency of driver mutations in *EGFR* (OR per doubling ancestry=1.1, p<0.0001), and a decreased frequency in *STK11* (OR=0.95, p<0.01) and *KRAS* (OR=0.95; p<0.0001), which remain associated after adjusting for smoking status (**Figure 2**). These associations were also observed when restricting to OncoKB actionable variants (Level 1, 2 and R1; Supplementary Figure 8). Additionally, we observed a positive association between *CDKN2A* SCNAs (OR=1.03; p=0.002), and a decrease in driver mutations in *BRAF* (OR=0.95; p=0.004) as EAS ancestry increases that persisted after adjustments for smoking. An association between EAS and increased driver mutations in *CTNNB1* was also observed (OR=1.04; p=0.009), which is attenuated by adjusting for multiple imputed smoking status (**Figure 2**), but not when adjusting for smoking in case-complete analysis (Supplemental Table 5, Supplemental Figure 7).

**Figure 2.**
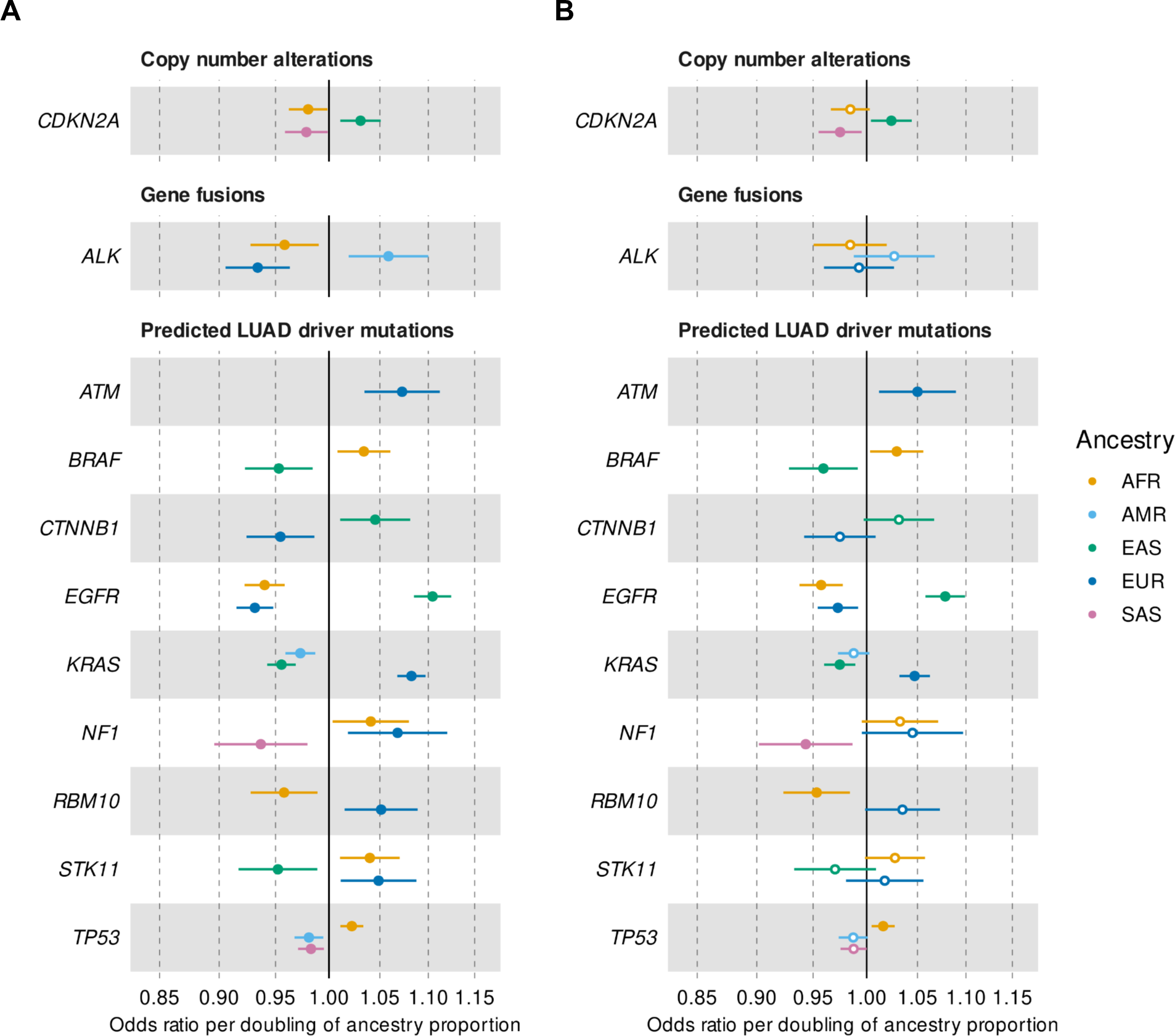
Forest plots for associations between genetic ancestry and SCNAs, gene fusions, and driver somatic mutations. The odds ratios (depicted as circles) and 95% confidence intervals (represented by horizontal lines) are shown for LUAD genes that met our criteria and displayed significant likelihood ratio tests (LRT) after adjusting for multiple testing at least in the univariable analysis. Colors represent ancestries as per color legend, full circles indicate significant logistic regression results, while empty circles denote cases where the odds ratio did not reach significance. Panel A, significant findings from univariable analyses without adjusting for smoking (n=13,196). Panel B, results after adjusting for smoking status derived from multiple imputation (n=13,196).

Associations were also identified between American Indigenous (AMR) ancestry, with a decreased frequency of driver mutations in *TP53* (OR=0.98; p=0.006) and *KRAS* (OR=0.97; p=0.002), although both become non-significant when controlling for smoking status (**Figure 2**). Previous studies have also shown an association between Native American ancestry and increased actionable variants in *EGFR*.^6^ Although we did not observe this association using boostDM predicted driver variants, we were able to replicate using OncoKB actionable variants (OR=1.04; p=0.0030), as well as with protein-altering variants in general (restricted to T/N cases; OR=0.97; p=0.001;Supplementary Figure 7). While the association of AMR with actionable variants in *EGFR* appears to be modulated by smoking, the association with protein altering variants remains significant after adjusting for smoking, either in the case-complete or multiple imputation analyses. Further, we find a significant association between AMR ancestry and *ALK* gene fusions (OR=1.06; p=0.003), which disappeared following adjustment for smoking (**Figure 2**).

For AFR ancestry, we observed decreased driver mutations in *RBM10* (OR=0.96), EGFR (OR=0.94) and fewer *ALK* fusions (OR=0.96) and *CDKN2A* SCNAs (OR=0.98), and increased driver mutations in *BRAF* (OR=1.04), *NF1* (OR=1.04), *STK11* (OR=1.03) and *TP53* (OR=1.02, all p<0.05; **Figure 2)**. All but *BRAF* and *RBM10* appear to be influenced by smoking (**Figure 2**). There were also associations with SAS ancestry showing decreased SCNAs in *CDKN2A* (OR=0.98) and driver mutations in *NF1* (OR=0.93) and *TP53* (OR=0.98, all p<0.05), with the latter being attenuated by smoking adjustment (**Figure 2**). Further, we observed associations between EUR ancestry and increased mutations in *KRAS* (1.08) and *ATM* (OR-1.07) that were not influenced by smoking, as well as increased driver mutations in *NF1* (OR=1.02), *RBM10* (OR= 1.05), and *STK11* (OR=1.05), and decreased mutations in *CTNNB1* (OR=0.95), and *ALK* fusions (OR=0.9; all p<0.05), which lost significance after adjusting for smoking status (**Figure 2**).

Finally, we also identified significant gene associations when considering any protein-altering variants regardless of their driver/actionable status and restricted to T/N cases only. These include associations between EAS and decreased number of variants in *KEAP1* (OR=0.94) and *SMARCA4* (OR=0.95), which appear to be independent of smoking status, while EUR showed the opposite effect (OR=1.05 and OR=1.05, correspondingly; all p<0.05), although attenuated by smoking (Supplementary Figure 8).

### Associations between imputed race and ethnicity categories and somatic mutations

In addition to the associations with genetic ancestry proportions, we conducted analyses with imputed R/E categories, using NH White as the reference group. Consequently, these analyses do not reveal associations specific to the NH White group. Results of all R/E LRT and association tests are given in Supplementary Table 6 and Files 4-6. Generally, we observed a strong concordance between AFR ancestry and NH Black, EAS and NH Asian, and AMR and Hispanic/Latino categories for associations with driver mutations in *BRAF*, *CTNNB1*, *EGFR*, *KRAS*, *STK11*, and *TP53*, as well as SCNAs in *CDKN2A* and *ALK* gene fusions (**Figure 3**). However, some discrepancies were noted; for instance, the associations between Hispanic/Latino and an increased or decreased number of driver mutations in *EGFR* and *KRAS*, respectively, were robust against MICE smoking adjustments. Notably absent were associations between Hispanic/Latino (seen with AMR) or NH Asian (seen with SAS) with mutations in *TP53*, and the association between SAS and decreased mutations in *NF1* did not emerge here (potentially because NH Asian conflates East and South Asians, with the latter being underrepresented in our cohort). Additionally, we found associations between Hispanic/Latino and an increased occurrence of *ALK* gene fusions (OR=2.9), and decreased driver mutations in *RBM10* (OR=0.43; both p<0.01). The analysis of OncoKB actionable and short protein-altering mutations essentially mirrors the results with driver mutations and genetic ancestry, with a few additions: associations between NH Black and increased protein altering mutations in *ALK* (OR=1.51) and *KEAP1* (OR=1.24); between Hispanic/Latinos and reduced mutations in *KEAP1* (OR=0.29) and *STK11* (OR=0.52); and between NH Asian and reduced mutations in *KEAP1* (OR=0.27) and *STK11* (OR=0.2; all p-value<0.3) Supplementary Figure 9).

**Figure 3.**
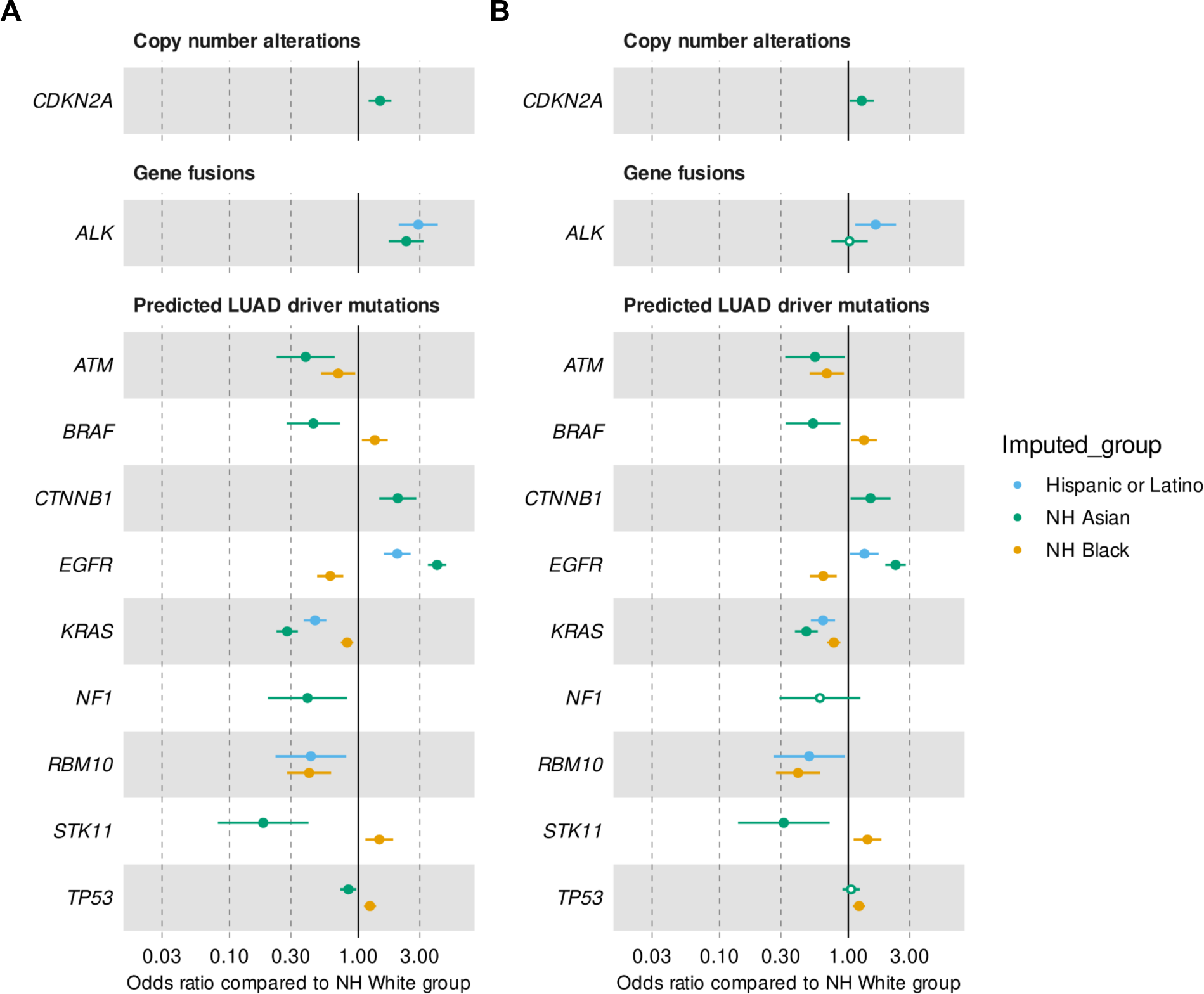
Forest plots for associations between imputed R/E categories and somatic mutations. Odds ratios (circles) and 95% confidence intervals (horizontal lines) for imputed race and ethnicity associations with somatic mutations using NH White as the reference group. LUAD genes that met our criteria and displayed significant likelihood ratio tests (LRT) after adjusting for multiple testing in at least in the univariable analysis are shown. Full circles indicate significant logistic regression results, while empty circles denote cases where the odds ratio did not reach significance. Panel A, all significant results from univariable analyses without adjustment for smoking N=13,103). Panel B, results in the multiple imputation analyses adjusted for smoking status (N=13,103).

### Distribution of mutations in CTNNB1 across imputed R/E categories

We aimed to further characterize the associations found with driver mutations in the β-Catenin gene, *CTNNB1*, and EAS and NH Asians. Initially, we observed that this association was identified using boostDM predicted drivers and total protein-altering mutations, and with both genetic ancestry and the imputed NH Asian category. Although the association is attenuated with smoking in multiple imputations for predicted driver mutations (see Table 2), the association with genetic ancestry remains unaffected after adjustment for smoking in protein-altering variants (Supplemental Figure 8), as well as for associations found with the imputed NH Asian category with predicted drivers (Figure 3) or protein-altering mutations (Supplemental Figures 8 and 9). Thus, our results suggest that smoking does not totally explain the increased burden of mutations in *CTNNB1* in EAS ancestry or NH Asian patients.

We examined the proportion of patients harboring a predicted driver mutation in *CTNNB1* across imputed R/E categories, distinguishing individuals with East Asian (EAS) and South Asian (SAS) ancestries within the NH Asian group. Figure 4 demonstrates that while the overall fraction of NH White patients in the cohort with such mutations is low (2.4%), albeit slightly higher in never-smoker NH White (3.6%). In contrast, this fraction is higher overall in NH Asian-EAS patients (5%; Fisher exact p-value<0.001) and even higher in never-smokers NH Asian-EAS patients (8.6%; p=0.001). It also appears in a considerable number of never-smoker Hispanic/Latino (3.8%) and NH Black (5.6%) patients (Supplementary Table 7).

**Figure 4.**
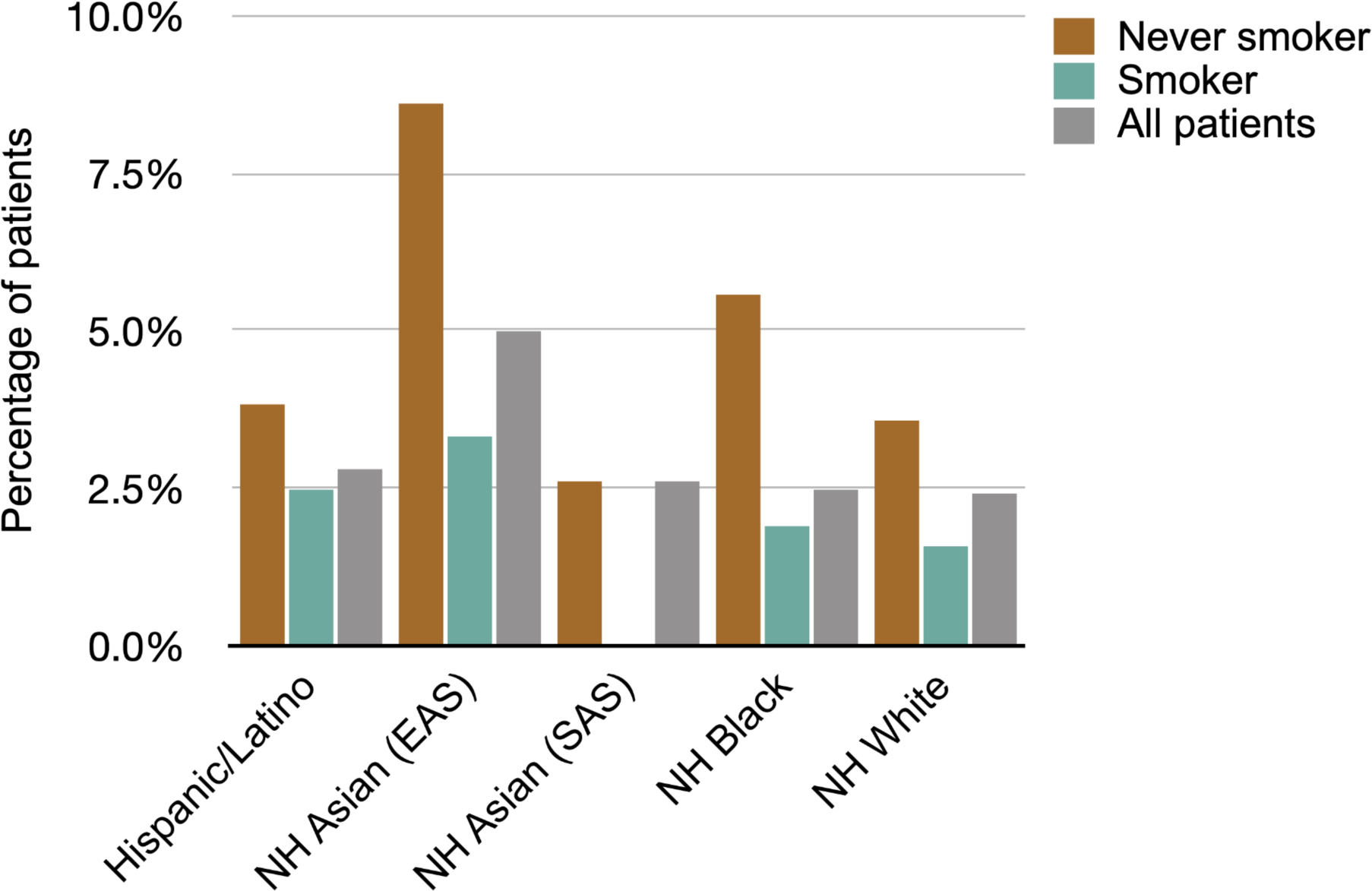
Distribution of driver mutations in *CTNNB1* by R/E and smoking status. Percentage of patients harboring a driver mutation in the *CTNNB1* gene predicted by boostDM, stratified by smoking status and R/E. Smoking status is color coded as per the legend. “Smoker” refers to “current or former smoker”. Only patients with measured smoking status were included in calculations. Imputed R/E categories are shown in the x-axis. In the case of the NH Asian category, we split patients by their proportions of EAS and SAS: EAS > SAS is indicated as NH Asian (EAS), whereas patients with SAS > EAS are designated as SAS. R/E is statistically significantly associated with the presence of mutations for all patients and never smoker cohorts (Fisher exact test, p=0.0018, and p=0.015, correspondingly). See Supplementary Table 7 for pair-wise statistical assessment of the differences.

## Discussion

In recent years, advances in genomic technologies have enabled a deeper understanding of the molecular underpinnings of lung cancer. However, the relationship between the molecular profiles of lung cancer and genetic ancestry or R/E categories remains relatively underexplored due to limited diversity in research studies. Previous studies in this area have been limited by a) a lack of diversity in research cohorts (e.g. TCGA^41^); b) smaller sample sizes; and c) reliance on US government-mandated R/E categories,^15^ which can be problematic. Considering these limitations, real-world clinical genomics databases represent a valuable resource for advancing our understanding of disparities and their molecular correlates in cancer research. Furthermore, the volume of data stored by such databases is expanding rapidly, ensuring that despite existing healthcare access disparities, minority groups are represented in substantially larger numbers compared to traditional research cohorts. Nevertheless, real-world data can have several shortcomings, notably substantial missingness in R/E data. The extent of this missingness is complex and varies across sources, ranging from 30-70%,^23^ but can largely be attributed to issues in data transmission and collection rather than simply patient abstention.^42,43^

The present study aimed to analyze data from a real-world clinico-genomic database to provide insights into the associations between lung cancer molecular profiles and genetic ancestry or R/E categories. To address the challenges typically associated with RWD, we implemented several strategies. Firstly, we inferred continental-level genetic ancestry from molecular data obtained during testing,^22^ allowing us to eschew categorical analysis in favor of logistic regression methods.^38^ We directly assessed the association between genetic ancestry proportions and somatic mutations of a specific type in LUAD genes via likelihood ratio tests (LRTs). P-values were examined if a positive association was identified. Compared to a strategy that forgoes LRTs and examines all logistic regression p-values directly, our approach reduces the multiple testing burden, ensuring high statistical power while minimizing the risk of type I errors. Additionally, when analyzing genetic ancestry proportions, it is essential to address the fact that they sum to one, resulting in collinearity (e.g., in admixed NH Black patients, it is common for AFR ancestry to increase as EUR ancestry decreases). We thus employed compositional logistic regression methods, applying an isometric log-ratio transform to include all proportions in the same model.^38^ This enabled us to identify associations with increased EUR ancestry, whereas in R/E analysis (see below), NH White is consistently used as the control, thereby not permitting the elucidation of these effects. To address the issue of missing clinical data, we utilized a multiple imputation strategy (MICE^39^) and performed an extensive assessment of the results to understand the accuracy of imputed values. This approach enabled us to increase power and to explore smoking’s role in our findings. For completeness, and because multiple imputation and complete case analysis are subject to different types of bias, we provided complete case analysis results from patients with known smoking status only (cf.

Supplementary Materials). Finally, for a categorical analysis involving R/E, we employed a previously developed ancestry-backed R/E imputation method demonstrated to be highly accurate within the Tempus data set.^33^

We identified associations between genetic ancestry or imputed R/E categories with different types of variants in LUAD-related genes. Our analysis included protein-altering short variants, SCNAs, rearrangements associated with gene fusions, actionable variants listed in OncoKB^37^ (Levels 1 & 2 and L1), and somatic driver variants for LUAD predicted by the boostDM algorithm.^35^ The boostDM algorithm evaluates and categorizes all possible single base substitutions in cancer genes according to their tumorigenic potential, drawing on a comprehensive analysis of mutations observed in a vast collection of sequenced tumors and annotating each site with relevant mutational features. This approach allowed us to compile a more comprehensive and potentially less biased list of driver mutations for genes for which boostDM models are available. Some of the associations we found with EAS, such as those with *CDKN2A* and *BRAF* persist after adjustment for smoking status. These associations are particularly noteworthy as the global decline in smoking has led to a higher prevalence of non-smoker LUAD, which is disproportionately represented in patients of East Asian ancestry.

One notable finding is the association between driver mutations in *CTNNB1* (the β-Catenin gene) and EAS ancestry. β-Catenin acts as a crucial co-activator in the oncogenic Wnt signaling pathway, where aberrations often lead to oncogenesis.^44^ Somatic mutations in *CTNNB1*, especially in exon 3, are implicated in this process, causing stabilization and accumulation of β-Catenin in cells and activating the Wnt/β-Catenin signaling pathway to increase cell proliferation.^45^ It has been reported that cancer cell lines with activating mutations in the *CTNNB1* gene are five times more sensitive to inhibitors of the spindle assembly checkpoint kinase (*TTK*), which are emerging as promising antineoplastic agents.^46^ Thus, *CTNNB1* mutations have been proposed as prognostic drug response biomarkers, potentially enabling the selection of patients most likely to benefit from *TTK* inhibitor therapy in proof-of-concept clinical trials.^44,47^ While *CTNNB1* mutations are prevalent in endometrial cancer and hepatocellular carcinoma,^46^ they have also been suggested as biomarkers for post-operative recurrence-free survival in *EGFR*-mutant LUAD.^48^ However, previous studies have indicated that *CTNNB1* mutations are rare in LUAD, with two large series reporting a frequency of approximately 2%—studies predominantly involving patients of European descent.^49,50^ In contrast, smaller case studies focusing on East Asian patients have indicated a higher frequency of *CTNNB1* mutations in LUAD.^51,52^ Our RWD study, which has a much larger sample size, now confirms the presence of *CTNNB1* driver mutations at appreciable frequencies in East Asian and Hispanic/Latino patients, as well as a statistically significant association with EAS ancestry. These results open the possibility of using *CTNNB1* mutations as biomarkers for the effectiveness of *TKK* inhibitors and prognosis in LUAD.

Our study’s methodology, robust in uncovering associations between genetic ancestry and mutational profiles in LUAD, faces several limitations worth noting. First, RWD may display ascertainment bias, as patients undergoing tumor profiling testing are likely those with later-stage cancer. This bias is exacerbated by disparities in insurance coverage and healthcare access, which may be confounded with R/E. Second, our approach to imputing R/E is limited to mutually exclusive categories, currently excluding Native American/Alaskan Natives—who are likely misclassified as “Hispanic/Latino”—and Hawaiian/Pacific Islanders, often misclassified as “NH Asian.” As the database expands to include more such patients, we plan to refine our methods to also impute these categories. Additionally, although multiple imputation is a vital tool for addressing missing data, it has its imperfections and potential biases. The directed acyclic graph in Supplementary Figure 1 highlights the roles of social determinants of health (SDOH), environmental, and genetic exposures in LUAD development, and considers how access to healthcare and screening affects cancer diagnosis, thus underscoring the complexity of these interacting factors. A significant limitation of our analysis is the lack of data on SDOH, which undeniably affects both the incidence and outcomes of lung cancer. This absence hinders our full understanding and integration of these factors into our analysis. Most importantly, it is essential to interpret our findings within the context that our methods establish associations rather than causality. This distinction necessitates cautious interpretation of our results, and underscores the importance of further research to clarify the intricate connections between genetic ancestry, mutational profiles, and lung cancer.

### Conclusion

Our methodology allowed us to identify both known and novel associations between somatic mutations and genetic ancestry or R/E groups. Our findings suggest that driver mutations in *CTNNB1* characterize a subgroup of mainly never-smoker LUAD patients more prevalent in East Asian populations, a potential biomarker of drug effectiveness. This result demonstrates how studies within diverse populations can aid the identification of new therapeutic approaches and provide insights that may ultimately help explain and address healthcare disparities.

## Data Availability

All data produced in the present work are contained in the manuscript.

## Acknowledgements

We would like to thank Yan Liu (Tempus), Carlos D. Bustamante and Alex Ioannidis (Stanford University) for statistical and methodology discussions and advice. We also acknowledge Rafael Esteller, Nick Rigan, and Arvind Prasad from the Tempus Lens team for their superb assistance in procuring de-identified data and correcting data problems needed for this work. We thank Frank Nothaft (formerly Tempus) for support with data access and Joel Dudley (formerly Tempus) for encouraging us to pursue this research. We thank Vanessa Nepomuceno from the Tempus Publications team for copy editing the manuscript.

## Funding

This study was funded by Tempus AI, Inc.

## Author Information

Authors and Affiliations

Tempus AI, Inc, Chicago, IL, USA

Brooke Rhead, Yannick Pouliot, Justin Guinney, and Francisco M. De La Vega

## Corresponding author

Francisco M. De La Vega.

## Contributions

Brooke Rhead: Methodology, data analysis, visualization, writing - review, editing. Yannick Pouliot: Data procurement, curation, analysis, methodology, writing - review, editing. Justin Guinney: Methodology, review, editing. Francisco De La Vega: Conceptualization, resources, supervision, writing - review, editing. All authors reviewed and suggested edits for the final version of the manuscript. The authors read and approved the final manuscript.

## Ethics declarations

Ethics approval and consent to participate.

All analyses were performed using de-identified data; The need for Institutional Review Board Approval was exempted by the IRB of Advarra, Inc., protocol no: Pro00042950, on April 15, 2020.

## Consent for publication

Not applicable.

## Competing Interests

B.R., Y.P., J. G., and F.M.D.L. are employees and have received stock options from Tempus AI, Inc.

---

**Supplementary Figure 1.**
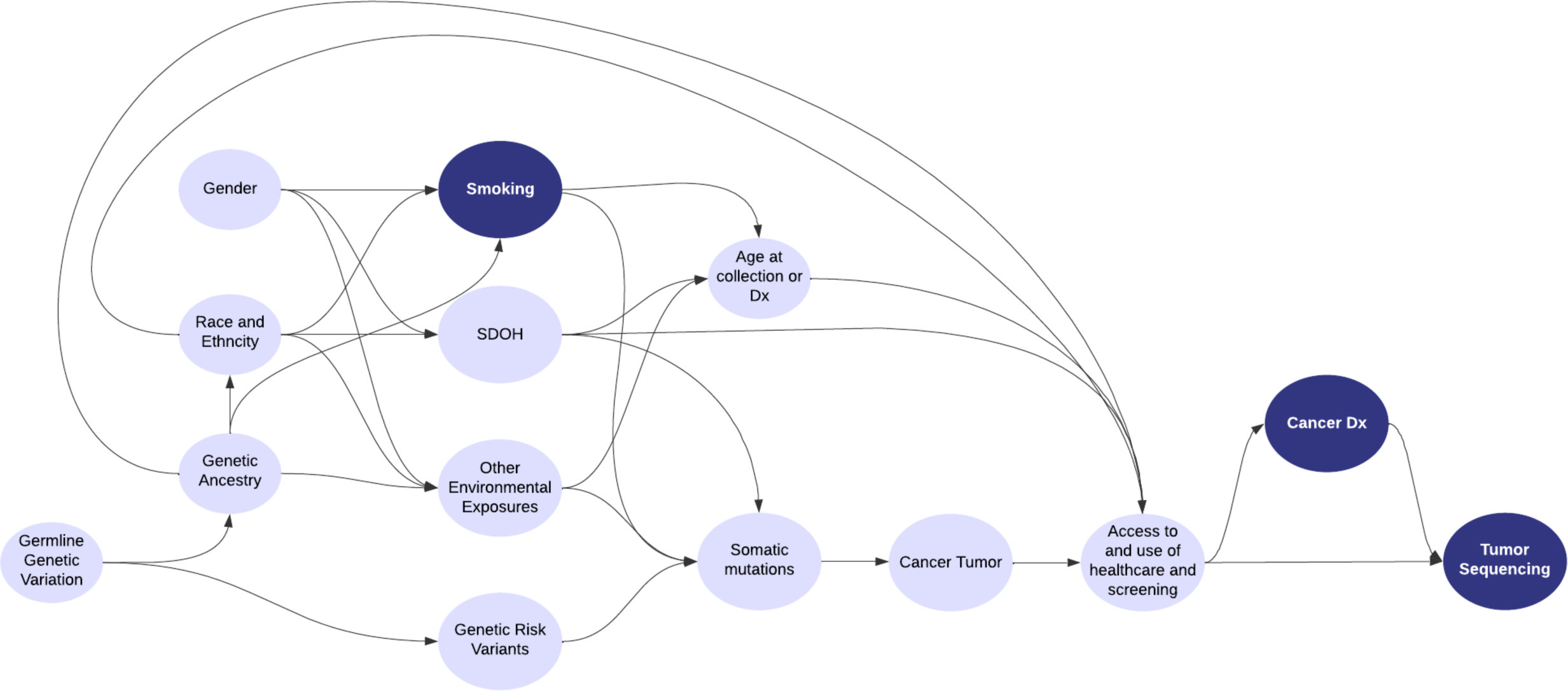
Directed Acyclic Graph (DAG) illustrating the causal pathways in lung adenocarcinoma progression. The graph delineates the relationships between race/ethnicity, genetic ancestry, gender, and key variables such as smoking, social determinants of health (SDOH), environmental and genetic exposures, and their collective impact on somatic mutations in genes. The downstream effects on cancer diagnosis, access to healthcare, and tumor sequencing are also represented. Dark blue shapes indicate variables that are controlled for in regression analysis or by selection into the study.

**Supplementary Figure 2.**
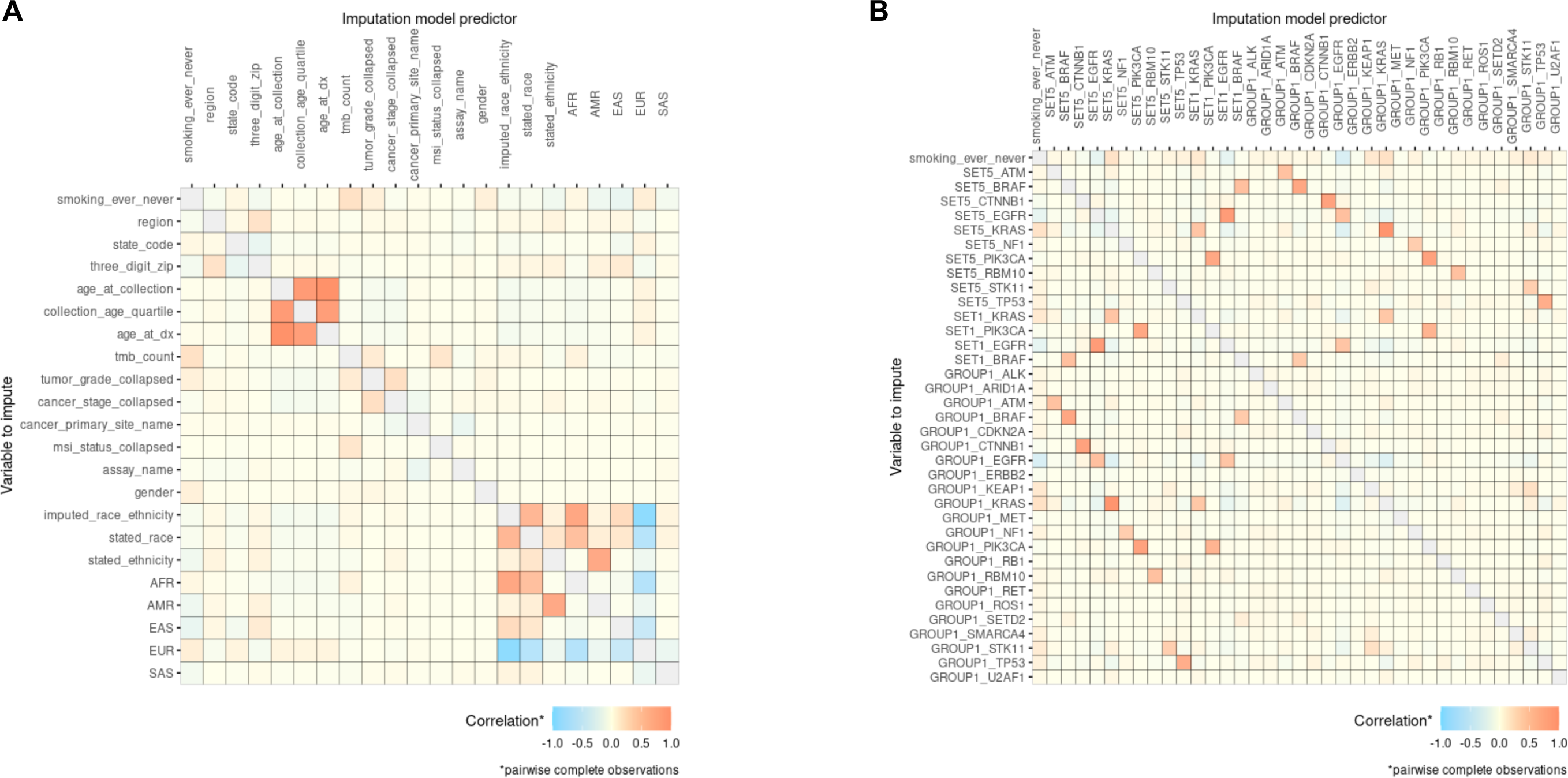
Correlation of clinical and somatic mutation data with smoking status. Panel A spearman correlation between clinical variables. Panel B spearman correlation between somatic mutation counts among genes and smoking status. Group 1, short protein altering variants; Group 2, SCNAs; Group 3, gene fusions, Set 1, OncoKb actionable variants; Set 5 BoostDM predicted drivers. Only cases with complete smoking status are included.

**Supplementary Figure 3.**
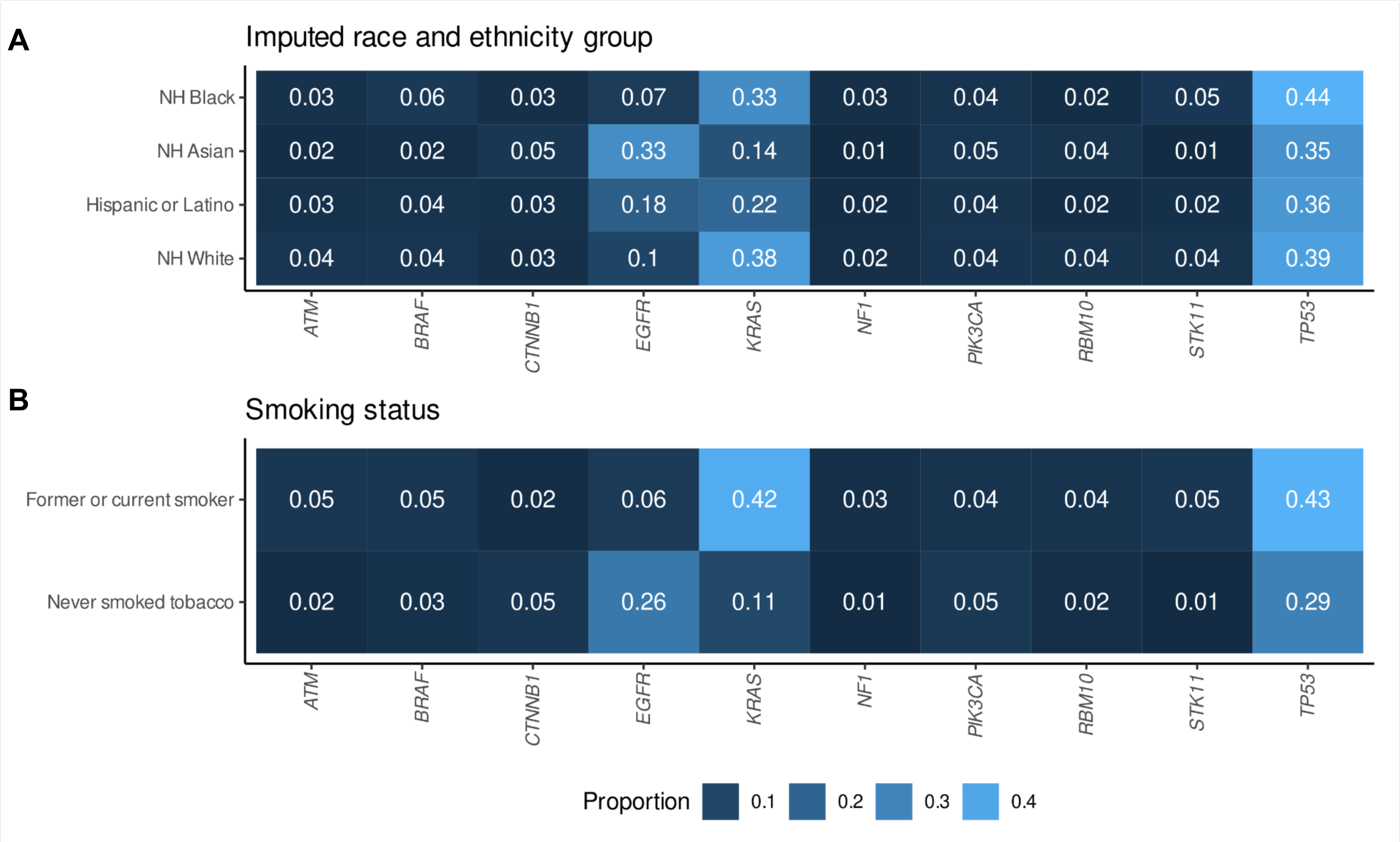
Proportion of patients with protein-altering variants in LUAD genes with respect to R/E and smoking status. Panel A: Fraction of patients carrying somatic protein-altering variants in selected LUAD genes (cf. Methods) with respect to imputed R/E categories. Panel B: raction of patients carrying somatic preten-latering variants in selected LUAD genes (cf. Methods) with respect to smoking status (only patients with available smoking status included).

**Supplementary Figure 4.**
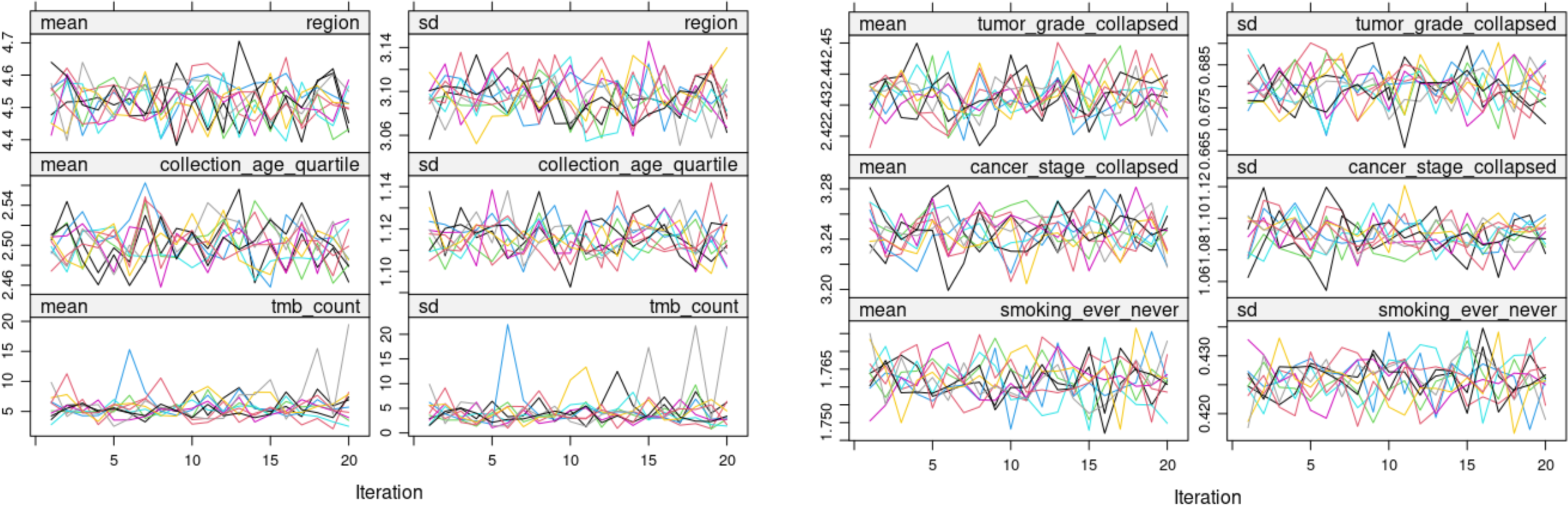
Convergence plots for multiple imputation. The mean and standard deviation (SD) for each of the imputed variables at each iteration of multiple imputation using MICE is shown. Variables included: Region, US geographical region of patient home address; Age at collection of tissue; TMB count (mutations/Mb); Tumor grade; Cancer stage; Smoking status, current (smoking), former smoker (ever), or never a smoker (never).

**Supplementary Figure 5.**
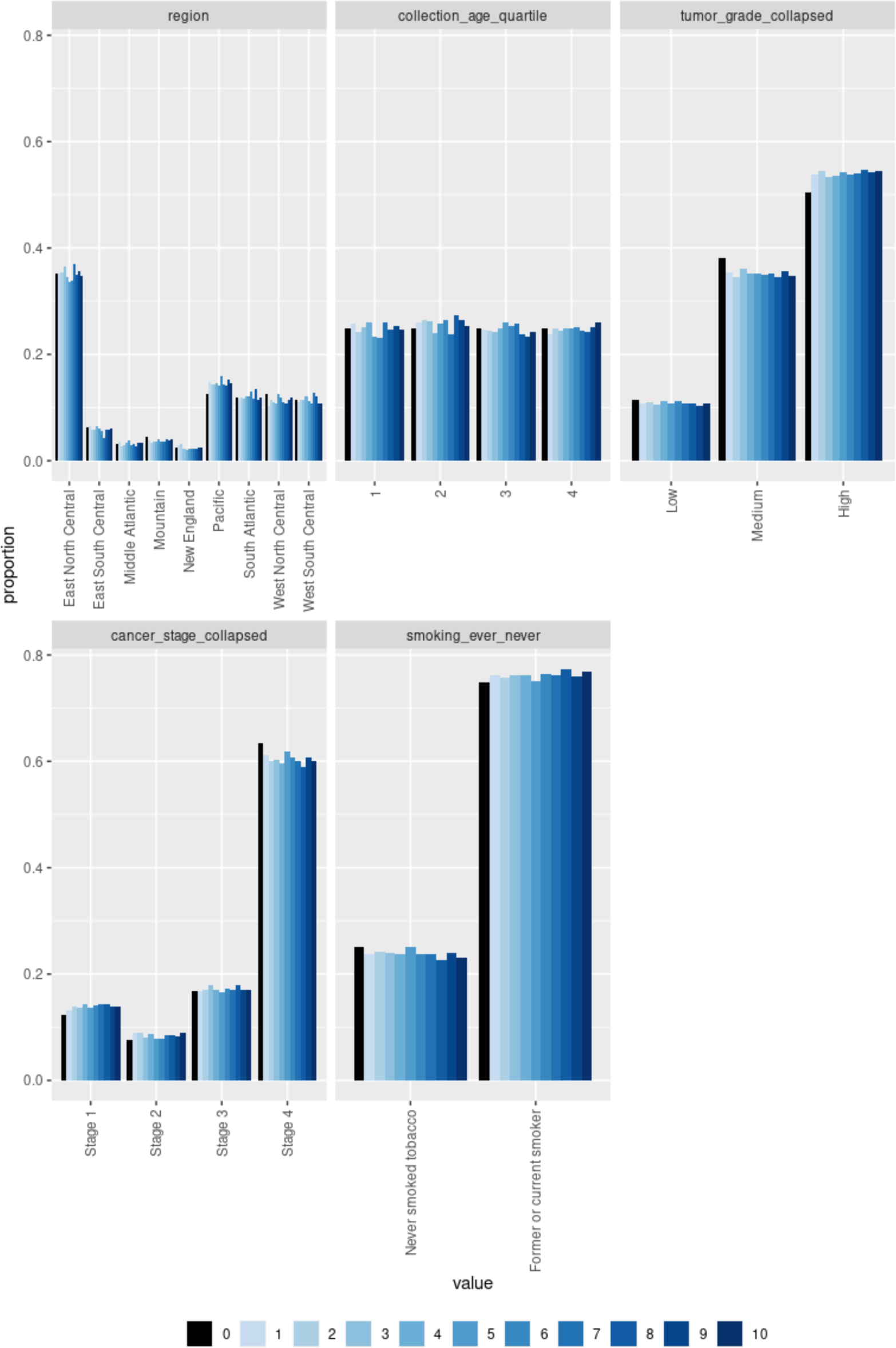
Distribution of values of multiply imputed categorical variables. The first bar (0) shows the distribution of measured categorical variables, remaining bars are data for each of the 10 imputed datasets (1-10). Regions: US geographical regions of patient home address.

**Supplementary Figure 6.**
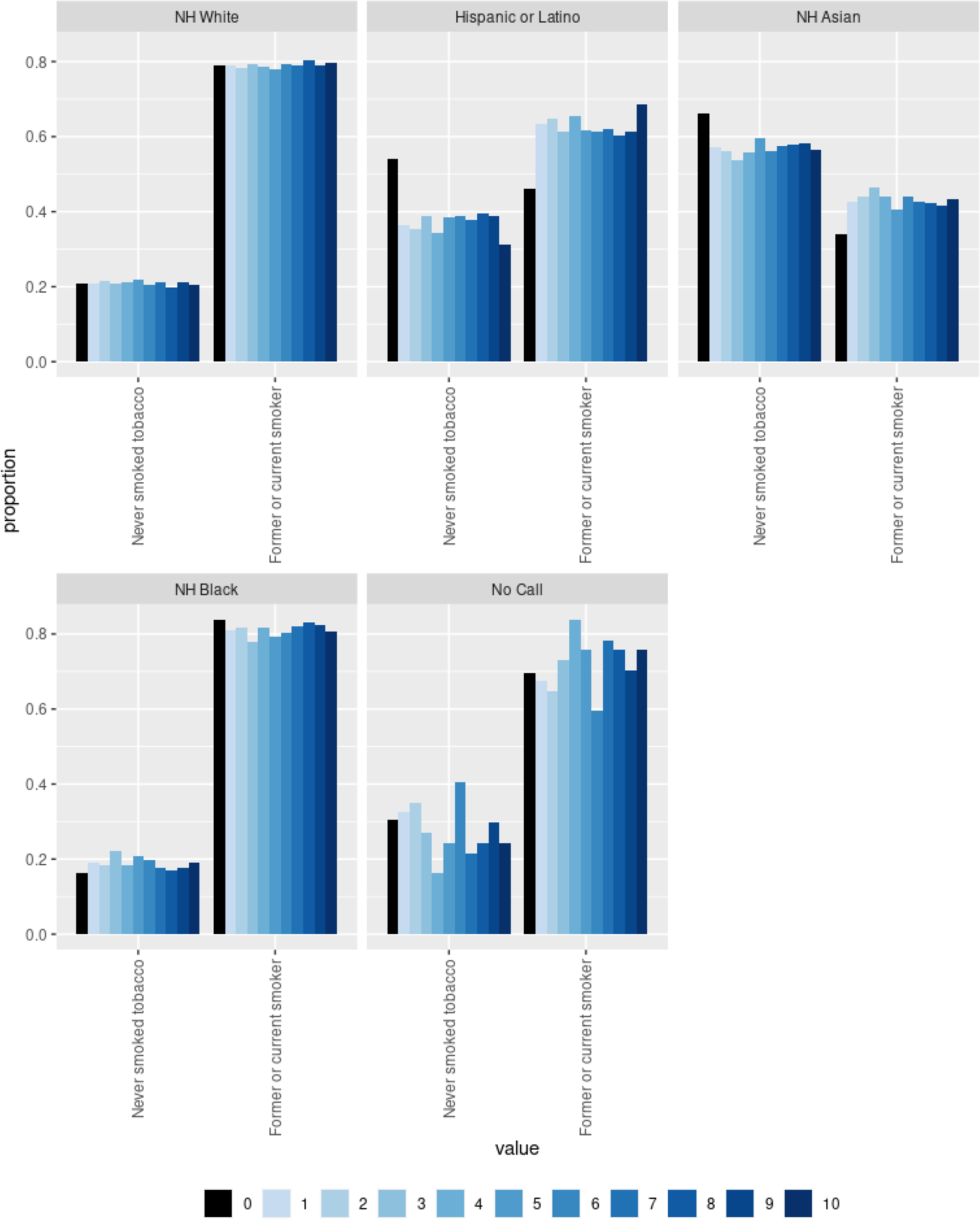
Distribution of multiply imputed smoking status by imputed race and ethnicity category. The first bar (0) shows the distribution of measured smoking status, subsequent bars show values for each of the 10 imputed datasets (1-10).

**Supplementary Figure 7.**
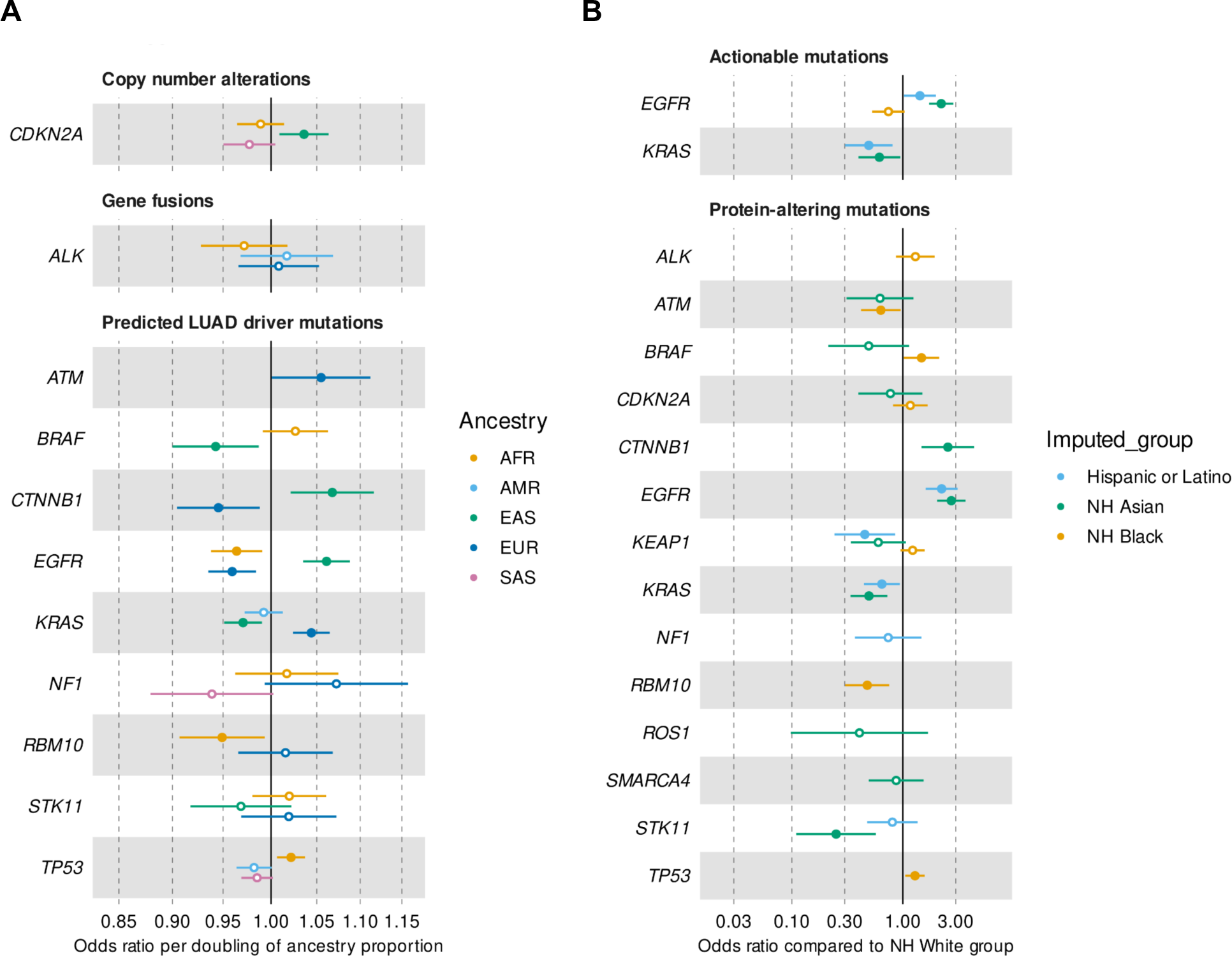
Forest plots for associations between genetic ancestry and imputed R/E with SCNAs, gene fusions, and driver somatic mutations adjusted for smoking for complete case analysis (n=7,133). The odds ratios (depicted as circles) and 95% confidence intervals (represented by horizontal lines) are shown for LUAD genes that met our criteria and displayed significant likelihood ratio tests (LRT) after adjusting for multiple testing at least in the univariable analysis. Colors representing ancestries as per color legend, full circles indicate significant logistic regression results, while empty circles denote cases where the odds ratio did not reach significance. Panel A, results for associations with genetic ancestry. Panel B, results for associations with imputed R/E categories using NH White as the reference group.

**Supplementary Figure 8.**
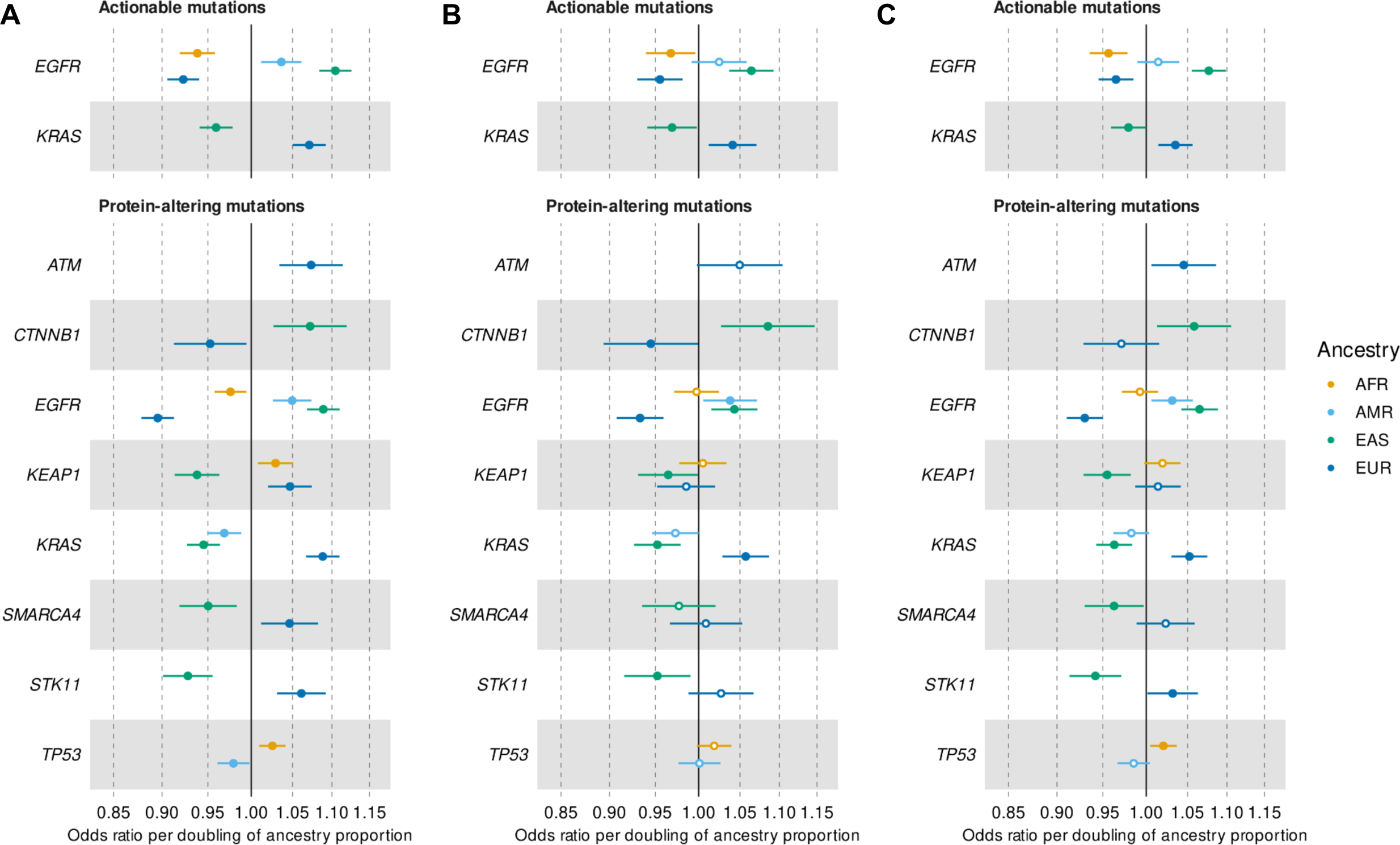
Forest plots for associations between genetic ancestry and actionable and protein-altering somatic mutations. The odds ratios (depicted as circles) and 95% confidence intervals (represented by horizontal lines) are shown for LUAD genes that met our criteria and displayed significant likelihood ratio tests (LRT) at least in the univariable analysis. Colors represent ancestries as per color legend, full circles indicate significant LRT results, while empty circles denote cases where the odds ratio did not reach significance after adjusting for multiple testing. Actionable mutations, OncoKB Level 1, 2, and R1. Panel A, significant findings from univariable analyses without adjusting for smoking (n=13,196). Panel B, results from complete case analyses adjusted for smoking status (n=7,133). Panel C, results after adjusted for smoking status derived from multiple imputation (n=13,196).

**Supplementary Figure 9.**
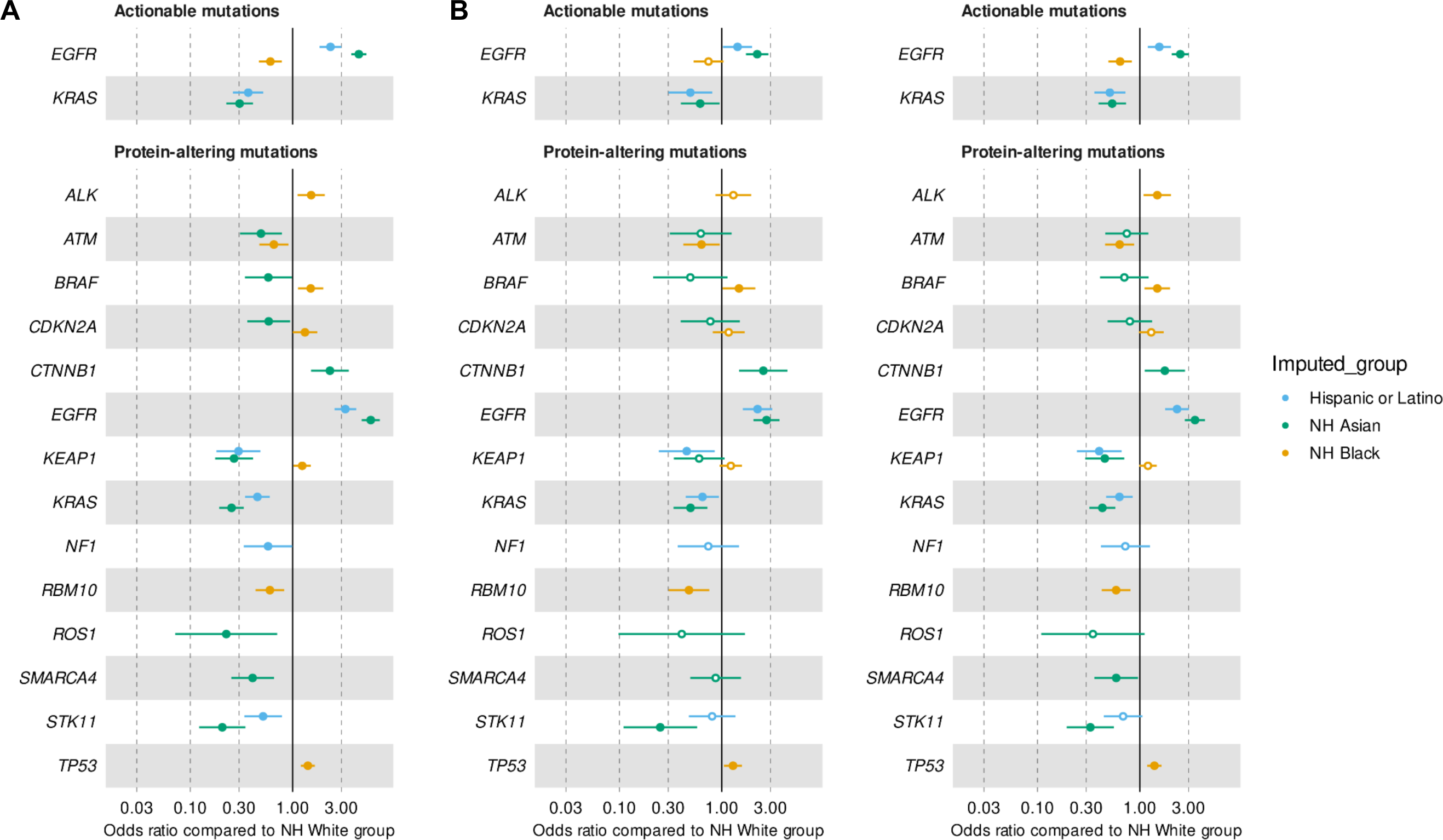
Forest plots for associations between imputed R/E categories and actionable and protein-altering somatic mutations. Odds ratios (circles) and 95% confidence intervals (horizontal lines) for imputed race and ethnicity associations with somatic mutations using NH White as the reference group. Full circles indicate significant LRT results, while empty circles denote cases where the odds ratio did not reach significance after adjusting for multiple testing. Actionable mutations, OncoKB Level 1, 2, and R1. Panel A, all significant results from univariable analyses without adjustment for smoking. Panel B, results in the complete case analyses adjusted for smoking status. Panel C, results in the multiple imputation analyses adjusted for smoking status.

**Supplementary Table 1.**
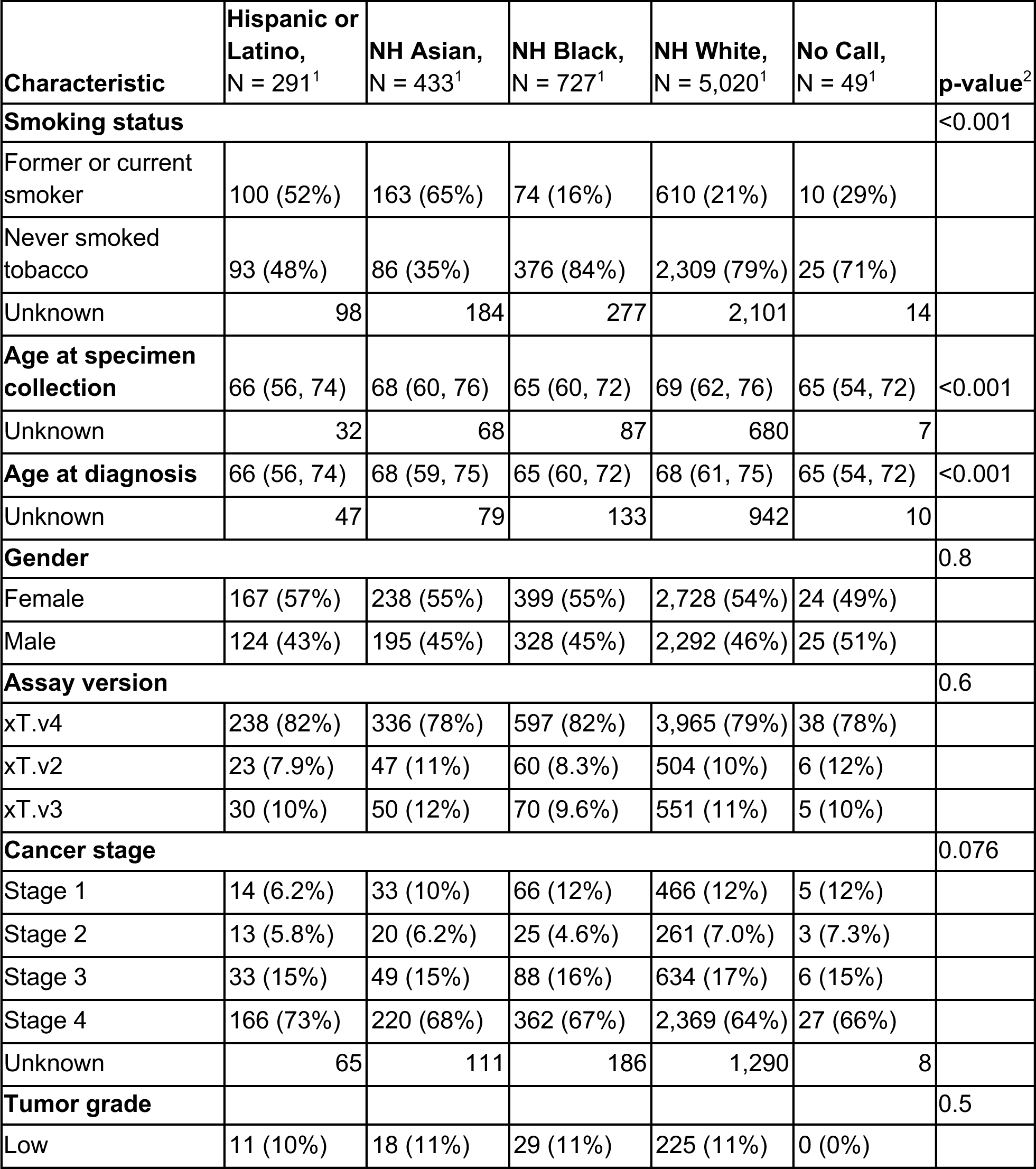
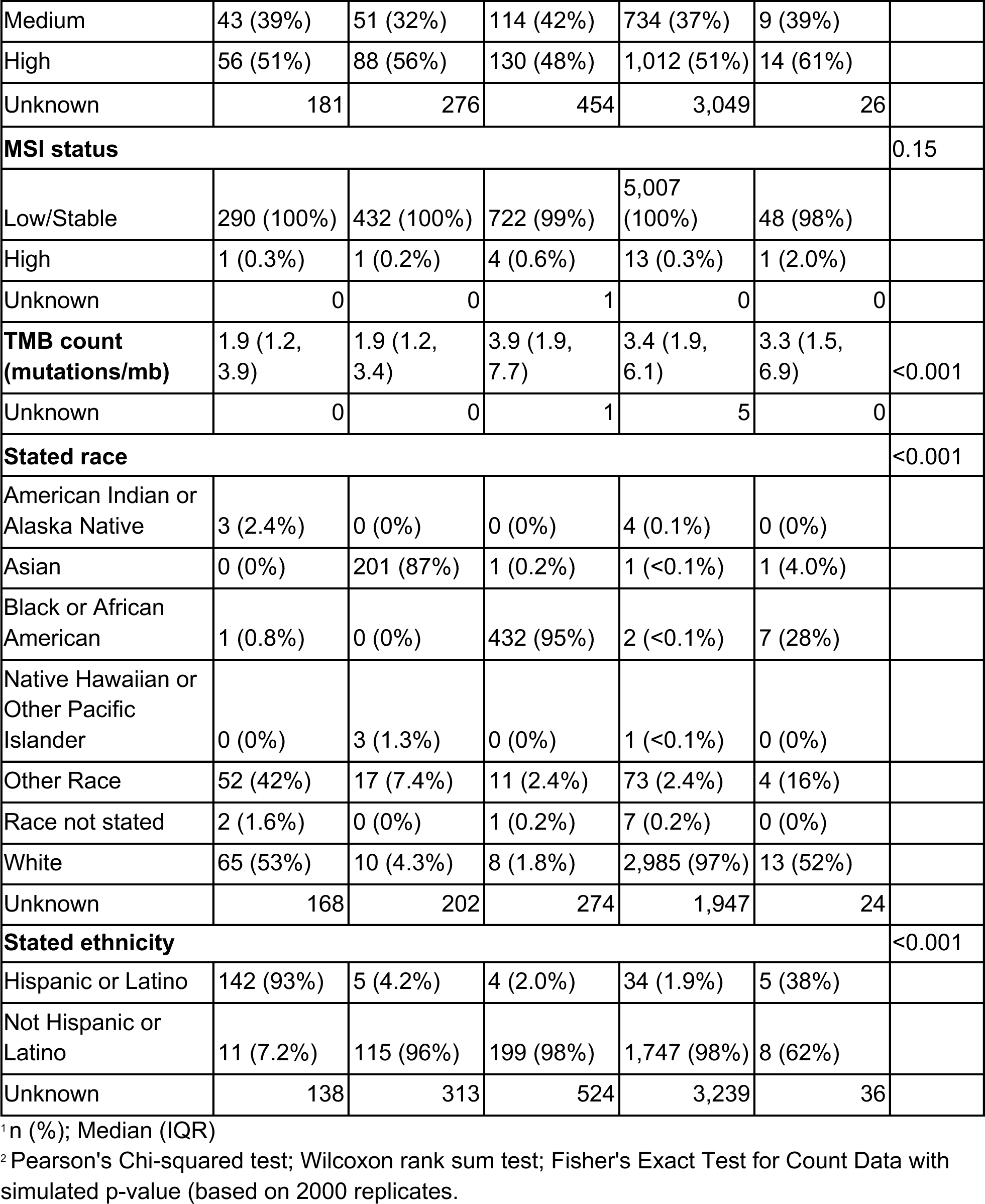
Characteristics of patients with tumor and normal samples by imputed race and ethnicity category. Last column shows the results of statistical tests (as defined in footnote) for differences of the characteristics by imputed race and ethnicity.

**Supplementary Table 2.** Cohort characteristics by smoking status.

| **Characteristic** | **Never smoked tobacco, N = 1,788<sup>1</sup>** | **Former or current smoker, N = 5,345<sup>1</sup>** | **p-value<sup>2</sup>** |
| --- | --- | --- | --- |
| **Imputed race and ethnicity category** |  |  | <0.001 |
| Hispanic or Latino | 184 (10%) | 157 (2.9%) |  |
| NH Asian | 317 (18%) | 162 (3.0%) |  |
| NH Black | 142 (7.9%) | 731 (14%) |  |
| NH White | 1,128 (63%) | 4,256 (80%) |  |
| No Call | 17 (1.0%) | 39 (0.7%) |  |
| **Age at specimen collection** | 68 (59, 76) | 67 (61, 74) | 0.3 |
| Unknown | 6 | 12 |  |
| **Age at diagnosis** | 67 (58, 76) | 67 (60, 74) | >0.9 |
| Unknown | 61 | 165 |  |
| **Gender** |  |  | <0.001 |
| Female | 1,222 (68%) | 2,766 (52%) |  |
| Male | 566 (32%) | 2,579 (48%) |  |
| **Assay version** |  |  | 0.3 |
| xT.v4 | 1,440 (81%) | 4,392 (82%) |  |
| xT.v2 | 160 (8.9%) | 452 (8.5%) |  |
| xT.v3 | 188 (11%) | 501 (9.4%) |  |
| **Cancer stage** |  |  | <0.001 |
| Stage 1 | 147 (9.9%) | 586 (13%) |  |
| Stage 2 | 109 (7.3%) | 333 (7.4%) |  |
| Stage 3 | 207 (14%) | 808 (18%) |  |
| Stage 4 | 1,027 (69%) | 2,794 (62%) |  |
| Unknown | 298 | 824 |  |
| **Tumor grade** |  |  | <0.001 |
| Low | 119 (16%) | 231 (9.4%) |  |
| Medium | 328 (45%) | 881 (36%) |  |
| High | 279 (38%) | 1,343 (55%) |  |
| Unknown | 1,062 | 2,890 |  |
| **MSI status** |  |  | 0.8 |
| Low/Stable | 1,780 (100%) | 5,325 (100%) |  |
| High | 7 (0.4%) | 19 (0.4%) |  |
| Unknown | 1 | 1 |  |
| **TMB count** | 2.3 (1.5, 3.9) | 5.0 (3.1, 8.3) | <0.001 |
| Unknown | 1 | 3 |  |
| **Tumor/normal tissue status** |  |  |  |
| Tumor and normal | 957 (54%) | 2,889 (54%) | 0.7 |
| Tumor only | 831 (46%) | 2,456 (46%) |  |
| **Stated race** |  |  | <0.001 |
| American Indian or Alaska Native | 0 (0%) | 10 (0.3%) |  |
| Asian | 185 (16%) | 85 (2.3%) |  |
| Black or African American | 94 (8.0%) | 521 (14%) |  |
| Native Hawaiian or Other Pacific Islander | 5 (0.4%) | 2 (<0.1%) |  |
| Other Race | 79 (6.7%) | 144 (3.9%) |  |
| Race not stated | 4 (0.3%) | 10 (0.3%) |  |
| White | 812 (69%) | 2,909 (79%) |  |
| Unknown | 609 | 1,664 |  |
| **Stated ethnicity** |  |  | <0.001 |
| Hispanic or Latino | 113 (15%) | 123 (5.3%) |  |
| Not Hispanic or Latino | 665 (85%) | 2,195 (95%) |  |
| Unknown | 1,010 | 3,027 |  |

<sup>1</sup> n (%); Median (IQR)
<sup>2</sup> Pearson's Chi-squared test; Wilcoxon rank sum test; Fisher's Exact Test for Count Data with simulated p-value (based on 2000 replicates)

**Supplementary Table 3.**
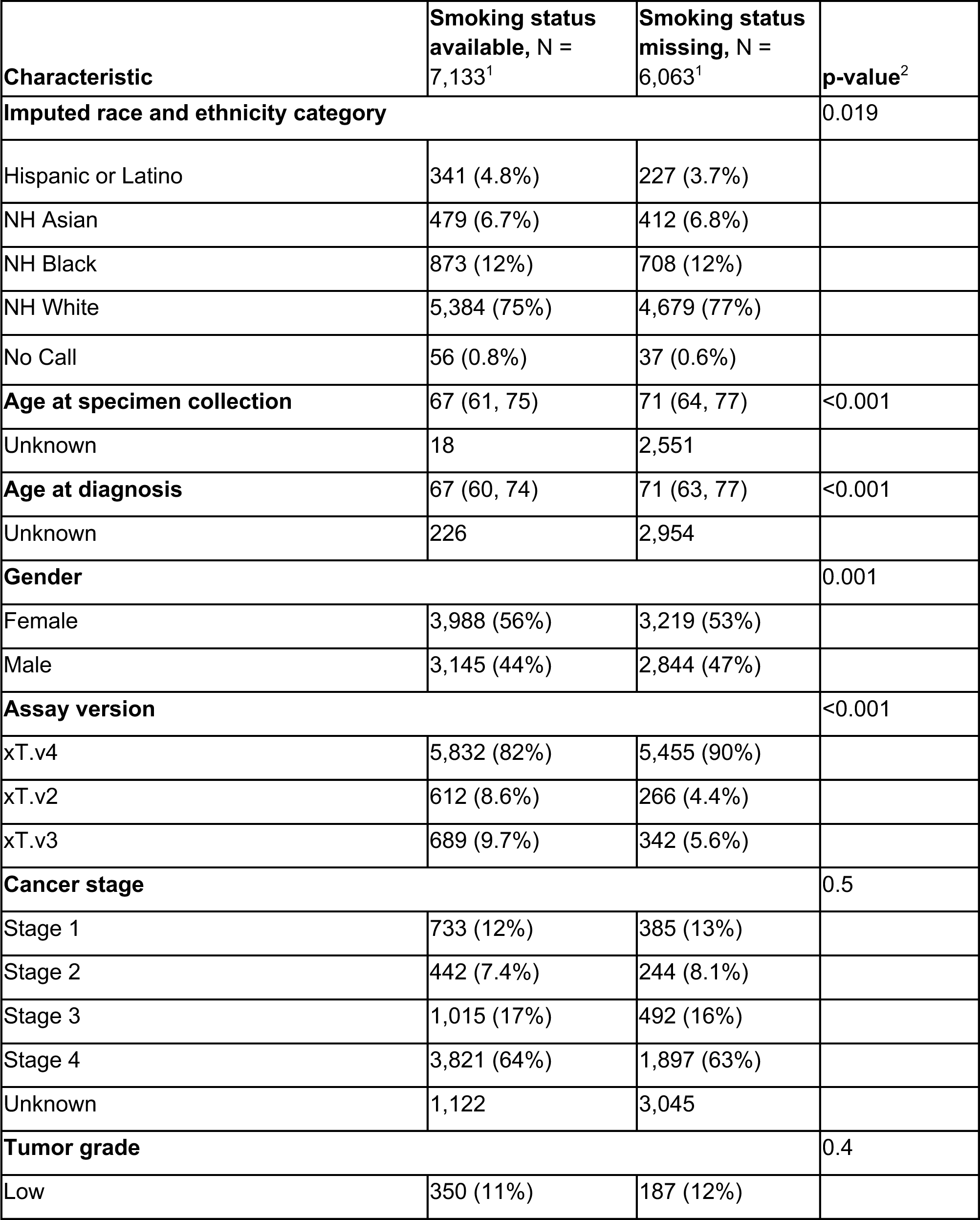
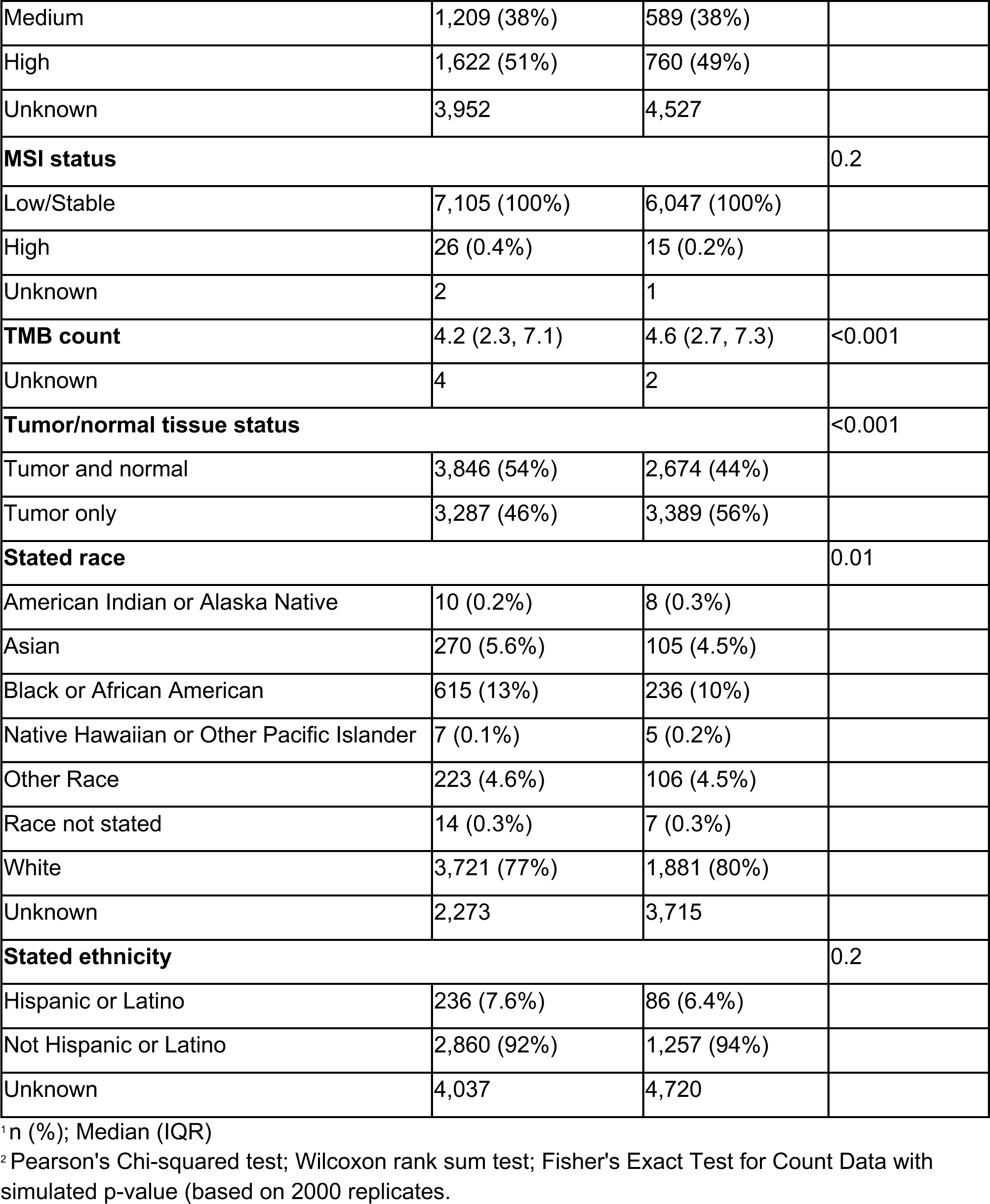
Cohort characteristics by availability of smoking status.

**Supplementary Table 4.** List of LUAD genes tested for association by mutation type. Genes tested varied depending on availability of data and type detected by assay (e.g. gene fusions and somatic copy number alterations, SCNAs) or available annotations (e.g. LUAD genes included in OncoKB as L1, L2 or R1, and available LUAD boostDM gene models).

| Mutation type | Number of genes | Genes |
| --- | --- | --- |
| Protein-altering<br>SNV/indels | 22 | *ALK, ARID1A, ATM, BRAF, CDKN2A, CTNNB1, EGFR, ERBB2, KEAP1, KRAS, MET, NF1, PIK3CA, RB1, RBM10, RET, ROS1, SETD2, SMARCA4, STK11, TP53, U2AF1* |
| SCNAs | 5 | *CDKN2A, EGFR, ERBB2, KRAS, MET* |
| Gene fusions | 1 | *ALK* |
| OncoKB actionable | 4 | *BRAF, EGFR, KRAS, PIK3CA* |
| boostDM predicted<br>drivers | 10 | *ATM, BRAF, CTNNB1, EGFR, KRAS, NF1, PIK3CA, RBM10, STK11, TP53* |

**Supplementary Table 5.** Summary of likelihood ratio test results for genetic ancestry and SCNA, gene fusion, and predicted driver mutations. Univariable and multiple imputation analyses were performed on N=13,196 patient samples, and complete case analyses were performed on N=7,133 patient samples. Bold indicates statistically significant corrected p-value.

| Mutation type | Gene | N tests | N (%) with mutation in all patients | N (%) with mutation in patients with known smoking status | Univariable analysis corrected p-value | Complete Case analysis corrected p-value | Multiple imputation analysis corrected p-value |
| --- | --- | --- | --- | --- | --- | --- | --- |
| SCNA | *CDKN2A* | 5 | 1,275 (10%) | 681 (10%) | **0.0017** | 0.2036 | **0.0319** |
| Fusion | *ALK* | 1 | 372 (3%) | 228 (3%) | **8.8e-06** | 0.7900 | 0.6876 |
| Driver | *ATM* | 10 | 503 (4%) | 258 (4%) | **0.0024** | 0.1862 | 0.0845 |
| Driver | *BRAF* | 10 | 579 (4%) | 305 (4%) | **0.0089** | 0.1469 | 0.0845 |
| Driver | *CTNNB1* | 10 | 347 (3%) | 175 (2%) | **0.0040** | **0.0074** | 0.2123 |
| Driver | *EGFR* | 10 | 1,248 (9%) | 661 (9%) | **5.47E-48** | **1.01E-07** | **1.08E-17** |
| Driver | *KRAS* | 10 | 4,569 (35%) | 2,411 (34%) | **1.57E-36** | **5.21E-05** | **1.57E-10** |
| Driver | *NF1* | 10 | 284 (2%) | 136 (2%) | **0.0024** | 0.1469 | 0.0686 |
| Driver | *RBM10* | 10 | 476 (4%) | 228 (3%) | **0.0062** | 0.1862 | **0.0296** |
| Driver | *STK11* | 10 | 461 (3%) | 252 (4%) | **0.0021** | 0.6165 | 0.2014 |
| Driver | *TP53* | 10 | 4,980 (38%) | 2,739 (38%) | **0.0006** | **0.0142** | **0.0029** |

**Supplementary Table 6.** Summary of likelihood ratio test results for imputed race and ethnicity and SCNA, gene fusion, and predicted driver mutations. Univariable and multiple imputation analyses were performed on N=13,103 patient samples, and complete case analyses were performed on N=7,077 patient samples. Bold indicates statistically significant corrected p-value.

| **Mutation type** | **Gene** | **N tests** | **N (%) with mutation in all patients** | **N (%) with mutation in patients with known smoking status** | **Univariable analysis corrected p-value** | **Complete Case analysis corrected p-value** | **Multiple imputation analysis corrected p-value** |
| --- | --- | --- | --- | --- | --- | --- | --- |
| SCNA | *CDKN2A* | 5 | 1264 (10%) | 675 (10%) | **0.0026** | 0.5944 | 0.5266 |
| Fusion | *ALK* | 1 | 368 (3%) | 225 (3%) | **1.24E-11** | 0.4187 | 0.1117 |
| Driver | *ATM* | 10 | 502 (4%) | 257 (4%) | **8.50E-05** | **0.0300** | **0.0118** |
| Driver | *BRAF* | 10 | 575 (4%) | 302 (4%) | **8.50E-05** | **0.0026** | **0.0118** |
| Driver | *CTNNB1* | 10 | 345 (3%) | 173 (2%) | **0.0022** | **0.0197** | 0.3224 |
| Driver | *EGFR* | 10 | 1242 (9%) | 657 (9%) | **1.39E-63** | **8.43E-09** | **1.13E-21** |
| Driver | *KRAS* | 10 | 4540 (35%) | 2394 (34%) | **1.87E-57** | **5.61E-06** | **1.33E-16** |
| Driver | *NF1* | 10 | 278 (2%) | 132 (2%) | **0.0238** | 0.0978 | 0.0999 |
| Driver | *RBM10* | 10 | 474 (4%) | 227 (3%) | **1.47E-06** | **0.0106** | **5.65E-06** |
| Driver | *STK11* | 10 | 461 (4%) | 252 (4%) | **4.29E-09** | **0.0197** | **9.27E-05** |
| Driver | *TP53* | 10 | 4943 (38%) | 2714 (38%) | **4.11E-05** | 0.0891 | **0.0236** |

**Supplementary Table 7.** Fisher exact test for differences in the presence of *CTNNB1* driver variants across R/E categories for all patients and stratified by current/former smokers or never smokers. 2-sided group test p-value under strata label; pairwise tests in cells (significant cells highlighted in orange). NH Asian patients were stratified by those with EAS or SAS ancestries.

|  | Hispanic or Latino | NH Asian (EAS) | NH Asian (SAS) | NH Black |
| --- | --- | --- | --- | --- |
| All patients (p=0.001836) |  |  |  |  |
| NH Asian (EAS) | 0.0534 | - | - | - |
| NH Asian (SAS) | 1 | 0.5727 | - | - |
| NH Black | 0.7588 | 0.0018 | 0.7187 | - |
| NH White | 0.5764 | 5.51 x 10<sup>-05</sup> | 0.7097 | 0.7938 |
| Current or former smoker (p=0.3619) |  |  |  |  |
| NH Asian (EAS) | 0.7453 | - | - | - |
| NH Asian (SAS) | 1 | 1 | - | - |
| NH Black | 0.5410 | 0.3481 | 1 | - |
| NH White | 0.3304 | 0.1062 | 1 | 0.5327 |
| Never smoker (p=0.01521) |  |  |  |  |
| NH Asian (EAS) | 0.0562 | - | - | - |
| NH Asian (SAS) | 1 | 0.3342 | - | - |
| NH Black | 0.4398 | 0.3338 | 0.6868 | - |
| NH White | 0.8338 | 0.0011 | 1 | 0.2458 |

## Notes

### Author Declarations

The Institutional Review Board of Advarra, Inc. waived ethical approval for this work.

